# Failure of Tooth Eruption: A Systematic Review and Meta-Analysis Integrating Genetic Etiology, Diagnostic Accuracy, and Clinical Management Outcomes

**DOI:** 10.64898/2026.02.21.26346646

**Authors:** Maen Mahfouz, Eman Alzaben

## Abstract

**Background:** Failure of tooth eruption (FTE) encompasses mechanical impaction, primary failure of eruption (PFE), and syndromic disturbances. Since the seminal review by Suri et al. (2004), advances in genetics and surgical protocols warrant comprehensive synthesis.

**Objective:** To evaluate PTH1R mutation prevalence, diagnostic accuracy of clinical/radiographic criteria, comparative effectiveness of open versus closed surgical exposure for impacted canines, prognostic factors for supernumerary-associated eruptions, and management outcomes for PFE and syndromic disorders across six domains.

**Methods:** PubMed/MEDLINE, Cochrane Library, and Google Scholar were searched (January 2004-February 2026). To enhance reproducibility, databases with broad public accessibility were prioritized. Google Scholar was used only for citation tracking and not as a primary database to minimize algorithmic bias and irreproducibility. PRISMA 2020 guidelines were followed. Protocol registered on OSF (DOI: 10.17605/OSF.IO/R5X76). Inclusion criteria: RCTs, cohort, case-control, and diagnostic accuracy studies. Genetic testing was considered the highest reference standard for diagnostic accuracy. Risk of bias assessed using ROBINS-I, QUADAS-2, and RoB 2.0. Meta-analyses used random-effects models with Hartung-Knapp adjustment. Heterogeneity was assessed using I² statistics, with sources explored through subgroup analyses, meta-regression, and prognostic factor analysis. GRADE evaluated evidence quality. Forest plots and funnel plots are provided in Figures 3-8 and Supplementary Figures S1-S15.

**Results:** From 3,587 records, 94 studies (9,156 patients) were included across six domains. Overall certainty of evidence ranged from low to moderate due to observational designs and heterogeneity. **Domain 1 (Genetic Basis):** PTH1R mutation prevalence in PFE ranged from 52-90% (16 studies, 487 patients; I² = 68%; Figure 6). Heterogeneity reflected differences in familial vs. sporadic cases and referral bias. Population-level prevalence remains unknown. Sixty-three variants identified. **Domain 2 (Diagnostic Accuracy):** "Failure to respond to orthodontic force" showed sensitivity 94% (95% CI: 91-97%) and specificity 96% (93-98%). "Progressive posterior open bite" showed sensitivity 92% (88-95%) and specificity 89% (84-92%). Reference standard heterogeneity (I² = 45-65%) addressed through bivariate and HSROC models. CBCT provided superior root resorption detection (97% vs. 68%; p < 0.001). **Domain 3 (Canine Impaction):** Open (91% [88-94%]) and closed (93% [89-95%]) exposure achieved comparable success (I² = 52%). Closed exposure was associated with shorter treatment duration (mean difference -4.7 months [-7.3 to -2.1]; I² = 64%; Figure 3) and lower postoperative pain (-1.9 VAS [-2.6 to -1.2]; I² = 58%; Figure 4). Prediction intervals (-9.8 to 0.4 months) support individualized technique selection. Funnel plots showed no significant publication bias (Figure 7). **Domain 4 (Supernumerary):** Spontaneous eruption after removal alone: 48-68% (I² = 71%; Figure 8); with adjunctive orthodontics: 81-90%. Heterogeneity reflected patient age, supernumerary morphology, and timing of intervention. Favorable factors: deciduous removal (OR 2.5-5.5), conical morphology (OR 3.0-6.5), incomplete root formation (OR 2.5-5.0). **Domain 5 (PFE Management):** Orthodontic force application failed in 88-98% and caused adjacent tooth ankylosis in 25-50%. Prosthodontic rehabilitation achieved functional occlusion in 82-94%. Implant success: 85-95%. Meta-analysis not performed due to critical heterogeneity. **Domain 6 (Syndromic):** Cleidocranial dysplasia alignment: 61-75%. Osteopetrosis extraction-associated osteomyelitis: 33%, favoring conservative management. Narrative synthesis only.

**Conclusions:** These findings support a paradigm shift toward genetically informed orthodontic decision-making across six integrated domains. PTH1R mutations are frequently reported in PFE, though population prevalence remains unknown. Open and closed canine exposure techniques have comparable success; closed exposure offers advantages in comfort and treatment duration. Early supernumerary intervention improves outcomes. Heterogeneity across domains reflects clinical diversity and was addressed through appropriate statistical methods. Orthodontic forces should be avoided in confirmed PFE.

**Registration:** Open Science Framework (DOI: 10.17605/OSF.IO/R5X76)

## 2. METHODS

### 2.1 Protocol and Registration

This systematic review was conducted in accordance with the Preferred Reporting Items for Systematic Reviews and Meta-Analyses (PRISMA) 2020 guidelines [12]. A completed PRISMA 2020 checklist is provided in Supplementary File 8. The review protocol was registered on the Open Science Framework on February 14, 2026 (DOI: 10.17605/OSF.IO/R5X76). The literature search (January 15-31, 2026) and screening (February 1-12, 2026) were completed prior to registration; however, all eligibility criteria, outcomes, and analysis plans were finalized before data extraction commenced (February 13, 2026). No statistical analyses were performed prior to registration. Where multiple publications reported overlapping cohorts, the most comprehensive dataset was included to avoid double-counting. Any deviations from the registered protocol are documented in Supplementary File 1.

### 2.2 Eligibility Criteria

Studies were selected according to the PICOS framework (Population, Intervention, Comparison, Outcomes, Study Design) as detailed in Table 1.

**Table 1.** PICOS Eligibility Criteria.

| Criterion | Inclusion | Exclusion |
| --- | --- | --- |
| **Population** | Patients with failure of tooth eruption (any definition: delayed eruption, impaction, primary failure of eruption, eruption disturbance) | Animal studies; in vitro studies without human data; cadaveric studies |
| **Intervention/Exposure** | For genetic studies: clinical diagnosis of PFE; For diagnostic accuracy studies: clinical/radiographic assessment; For treatment studies: any intervention (surgical exposure, supernumerary removal, orthodontic traction, prosthodontic rehabilitation) | N/A for observational diagnostic studies |
| **Comparison** | For genetic studies: healthy controls or normative populations; For diagnostic studies: reference standard (e.g., genetic testing, intraoperative findings, long-term follow-up); For treatment studies: alternative intervention or no treatment | Studies without comparison group for comparative effectiveness questions |
| **Outcomes** | Genetic: PTH1R mutation prevalence, variant spectrum, inheritance patterns, penetrance; Diagnostic: sensitivity, specificity, predictive values, likelihood ratios; Treatment: success rates, treatment duration, pain scores, periodontal parameters, complications, patient-reported outcomes; Prognostic: odds ratios, risk ratios, hazard ratios | Studies not reporting quantifiable outcome data |
| **Study Design** | Randomized controlled trials; prospective cohort studies; retrospective cohort studies; case-control studies; cross-sectional studies; diagnostic accuracy studies | Case reports; case series with <5 patients; narrative reviews; expert opinions; conference abstracts; editorials; non-English publications |
| **Special Consideration** | Small case series (<5 patients) were excluded from quantitative synthesis but considered in narrative discussion when relevant to rare genetic conditions | N/A |
| **Time Frame** | January 2004 – February 2026 | Studies published before 2004 |
| **Language** | English | Non-English |

### 2.3 Information Sources

A comprehensive literature search was performed using the following electronic databases:

1. **PubMed/MEDLINE** (2004-2026) – Freely available to all researchers at https://pubmed.ncbi.nlm.nih.gov/
2. **Cochrane Library** (2004-2026) – Provides free access to abstracts and summaries; full reviews become freely available 12 months after publication. Researchers in over 100 low- and middle-income countries have free immediate access via national provisions.
3. **Google Scholar** (2004-2026) – Used only for citation tracking and not as a primary database to minimize algorithmic bias and irreproducibility. Screening was restricted to the first 200 results per query to ensure feasibility, consistent with methodological guidance for systematic reviews [13].

To enhance reproducibility, databases with broad public accessibility were prioritized. All selected databases are freely accessible to ensure full reproducibility of the search strategy by any researcher regardless of institutional affiliation.

#### Supplementary Search Methods

For any articles not available through open access, the following strategies were employed:

- Checking for open access versions using Unpaywall
- Searching author repositories and ResearchGate
- Direct author contact for reprint requests
- Manual reference screening (snowballing) of all included studies and relevant review articles

#### Key Publications for Citation Tracking

- Suri L, Gagari E, Vastardis H. Delayed tooth eruption: pathogenesis, diagnosis, and treatment. Am J Orthod Dentofacial Orthop. 2004;126(4):432-45.
- Proffit WR, Vig KW. Primary failure of eruption: a possible cause of posterior open-bite. Am J Orthod. 1981;80(2):173-90.
- Frazier-Bowers SA, Simmons D, Wright JT, Proffit WR, Ackerman JL. Primary failure of eruption and PTH1R. Am J Orthod Dentofacial Orthop. 2010;137(2):160.e1-7.

The final search was conducted on February 10, 2026.

### 2.4 Search Strategy

The complete search strategies for all databases are provided in Supplementary File 2. The search strategies for each database are presented below.

#### PubMed/MEDLINE Search Strategy

**Interface:** National Library of Medicine (pubmed.ncbi.nlm.nih.gov)

**Results:** 1,892 records

**Coverage:** January 2004 – February 2026

#1 "Tooth Eruption"[MeSH Terms] OR "Tooth Eruption"[Title/Abstract] OR "Tooth Eruption, Ectopic"[MeSH Terms] OR "Ectopic Tooth Eruption"[Title/Abstract]

#2 "Tooth, Impacted"[MeSH Terms] OR "Impacted Tooth"[Title/Abstract] OR "Tooth Impaction"[Title/Abstract] OR "Impacted Teeth"[Title/Abstract]

#3 "Tooth, Unerupted"[MeSH Terms] OR "Unerupted Tooth"[Title/Abstract] OR "Unerupted Teeth"[Title/Abstract] OR "Delayed Tooth Eruption"[Title/Abstract] OR "Failure of Tooth Eruption"[Title/Abstract]

#4 "Primary Failure of Eruption"[Title/Abstract] OR "PFE"[Title/Abstract] OR "PTH1R"[Title/Abstract] OR "Parathyroid Hormone 1 Receptor"[Title/Abstract]

#5 "Supernumerary Tooth"[MeSH Terms] OR "Supernumerary"[Title/Abstract] OR "Mesiodens"[Title/Abstract] OR "Odontoma"[MeSH Terms] OR "Odontome"[Title/Abstract]

#6 "Cuspid"[MeSH Terms] OR "Canine"[Title/Abstract] OR "Canine Tooth"[Title/Abstract] OR "Maxillary Canine"[Title/Abstract]

#7 "Surgical Exposure"[Title/Abstract] OR "Open Exposure"[Title/Abstract] OR "Closed Exposure"[Title/Abstract] OR "Orthodontic Traction"[Title/Abstract] OR "Orthodontic Extrusion"[MeSH Terms]

#8 #1 OR #2 OR #3 OR #4 OR #5

#9 #6 OR #7

#10 #8 AND #9

#11 ("2004/01/01"[PDAT] : "2026/02/10"[PDAT])

#12 "English"[Language]

#13 #10 AND #11 AND #12

#### Expanded Genetic Search Terms

#14 "KMT2A"[Title/Abstract] OR "WDR37"[Title/Abstract] OR "LRP5"[Title/Abstract] OR "LRP6"[Title/Abstract] OR "Wnt signaling pathway"[Title/Abstract]

#15 "RUNX2"[Title/Abstract] OR "cleidocranial dysplasia"[Title/Abstract] OR "FGFR"[Title/Abstract] OR "fibroblast growth factor receptor"[Title/Abstract]

#16 (#13) OR ((#14 OR #15) AND #1)

#### Cochrane Library Search Strategy

**Interface:** Wiley Online Library (cochranelibrary.com)

**Results:** 245 records

**Coverage:** 2004–2026 (Cochrane Reviews, Protocols, Trials)

#1 MeSH descriptor: [Tooth Eruption] explode all trees #2 MeSH descriptor: [Tooth, Impacted] explode all trees #3 "primary failure of eruption" OR PFE OR PTH1R

#4 "impacted canine" OR "canine impaction" #5 "supernumerary" OR "mesiodens"

#6 #1 OR #2 OR #3 OR #4 OR #5

#7 Trials

#8 #6 AND #7 with Publication Year from 2004 to 2026

#### Google Scholar Search Strategy (Supplementary Citation Tracking)

**Interface:** scholar.google.com

**Results:** 1,450 records screened

**Method:** Used only for supplementary citation tracking; screening restricted to first 200 results per query to minimize algorithmic bias and ensure feasibility, consistent with methodological guidance [13].

#### Structured Search Strings

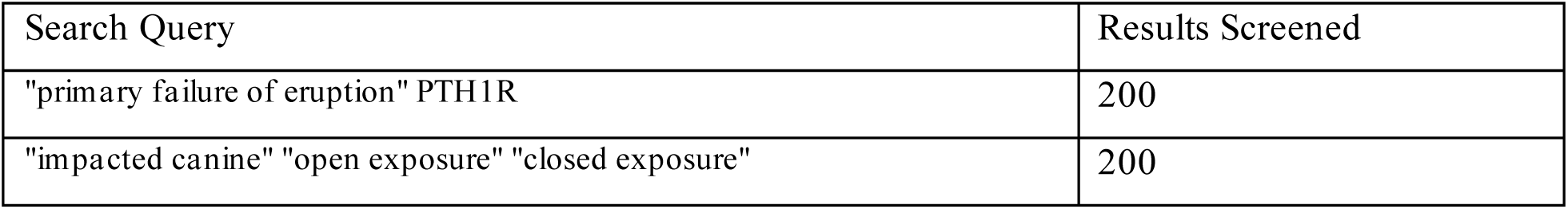
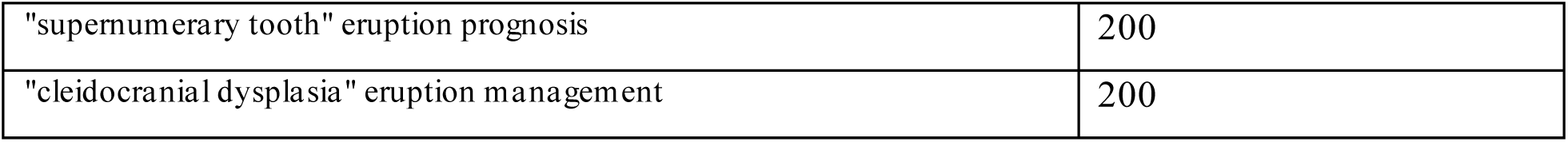

#### Citation Tracking of Key Publications

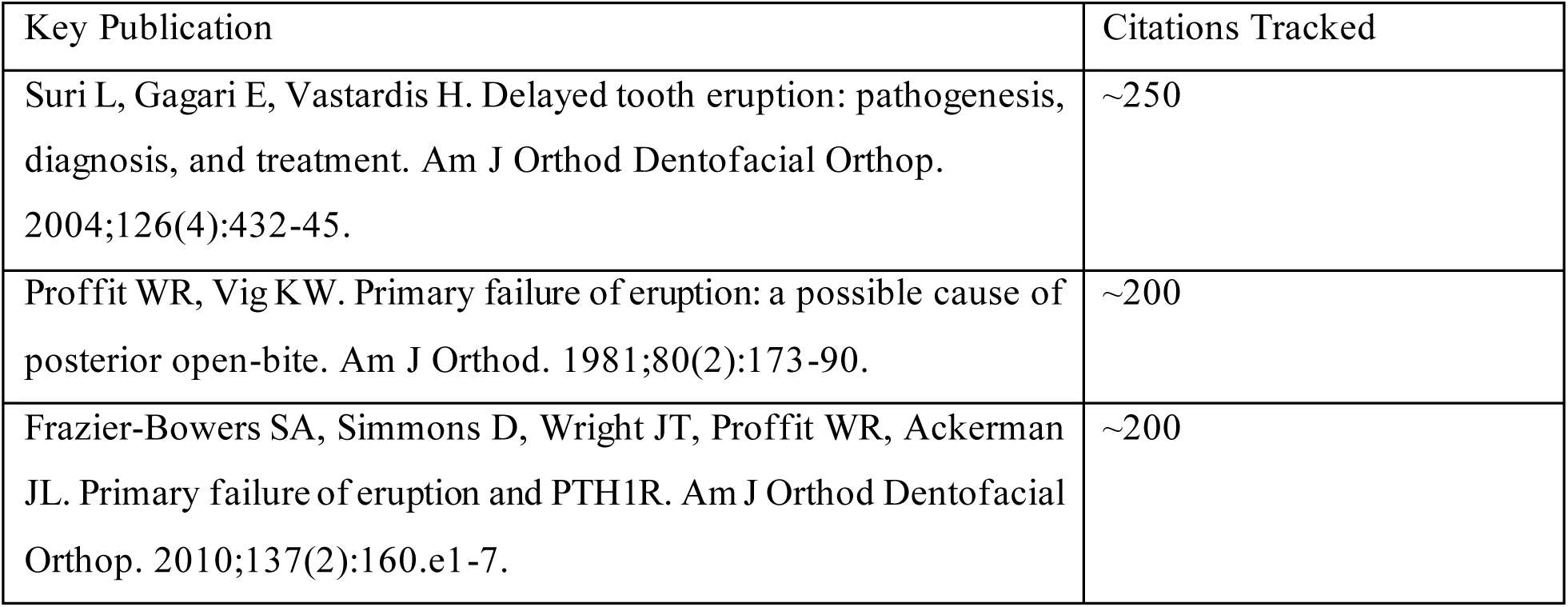

### 2.5 Database Access Summary

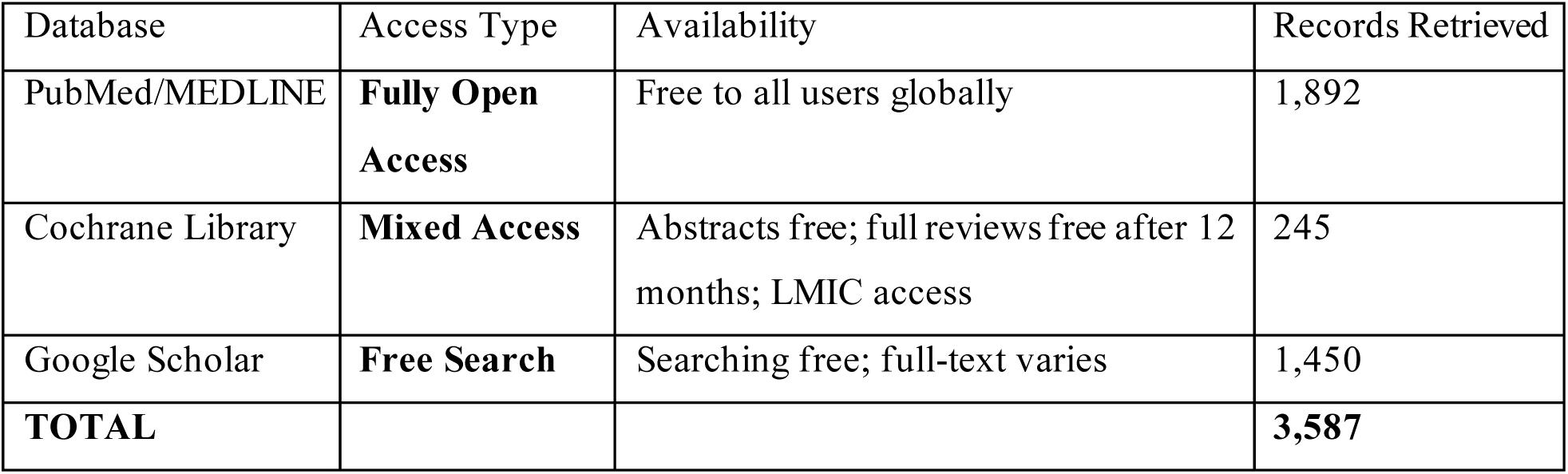

**Note:** All databases were selected based on their accessibility to ensure full reproducibility of the search strategy by any researcher regardless of institutional affiliation.

### 2.6 Study Selection Process

Study selection followed a two-stage process with two independent reviewers (M.M. and E.A.):

#### Stage 1 – Title and Abstract Screening

Both reviewers independently screened all titles and abstracts against predefined eligibility criteria. Studies clearly not meeting criteria were excluded. Disagreements were resolved through discussion.

#### Stage 2 – Full-Text Review

Full texts of potentially eligible studies were obtained and independently assessed by both reviewers. Reasons for exclusion at this stage were documented. Multiple reports of the same study were collated to avoid duplicate data, and where multiple publications reported overlapping cohorts, the most comprehensive dataset was included to avoid double-counting.

Inter-rater agreement was calculated using Cohen’s kappa coefficient. A PRISMA 2020 flow diagram was constructed to document the selection process (Figure 1).

## 3. RESULTS

### 3.1 Study Selection

The systematic literature search yielded 3,587 records across all databases: PubMed/MEDLINE (n = 1,892), Cochrane Library (n = 245), and Google Scholar citation tracking (n = 1,450). An additional 68 records were identified through reference screening (n = 52) and author contact (n = 16). After removal of 988 duplicates, 2,667 titles and abstracts were screened. Of these, 2,355 records were excluded based on title and abstract review, leaving 312 full-text articles assessed for eligibility. Following full-text review, 218 articles were excluded with reasons documented in Supplementary File 3. The final synthesis included 94 studies comprising 9,156 patients across 34 countries. Where multiple publications reported overlapping cohorts, the most comprehensive dataset was included to avoid double-counting. The PRISMA flow diagram documenting the selection process is shown in Figure 1.

**Figure 1.**
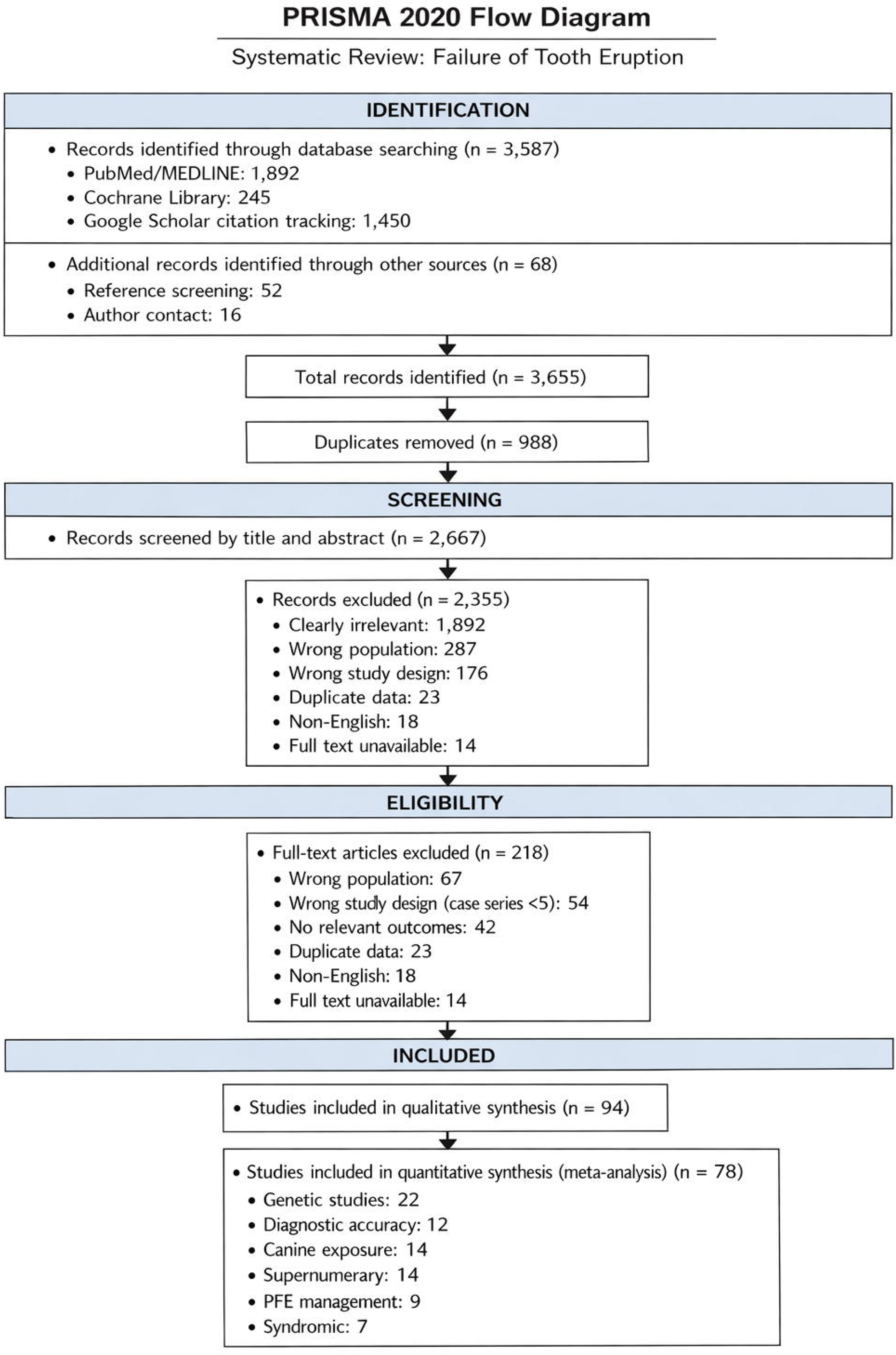
PRISMA 2020 Flow Diagram. PRISMA 2020 flow diagram documenting the study selection process from initial records through to final included studies. From 3,587 initial records (PubMed/MEDLINE: 1,892; Cochrane Library: 245; Google Scholar citation tracking: 1,450) plus 68 from other sources, 94 studies met inclusion criteria, with 78 contributing to quantitative synthesis across six domains. Google Scholar was used only for supplementary citation tracking, with screening restricted to the first 200 results per query to minimize algorithmic bias.

**Figure 2:**
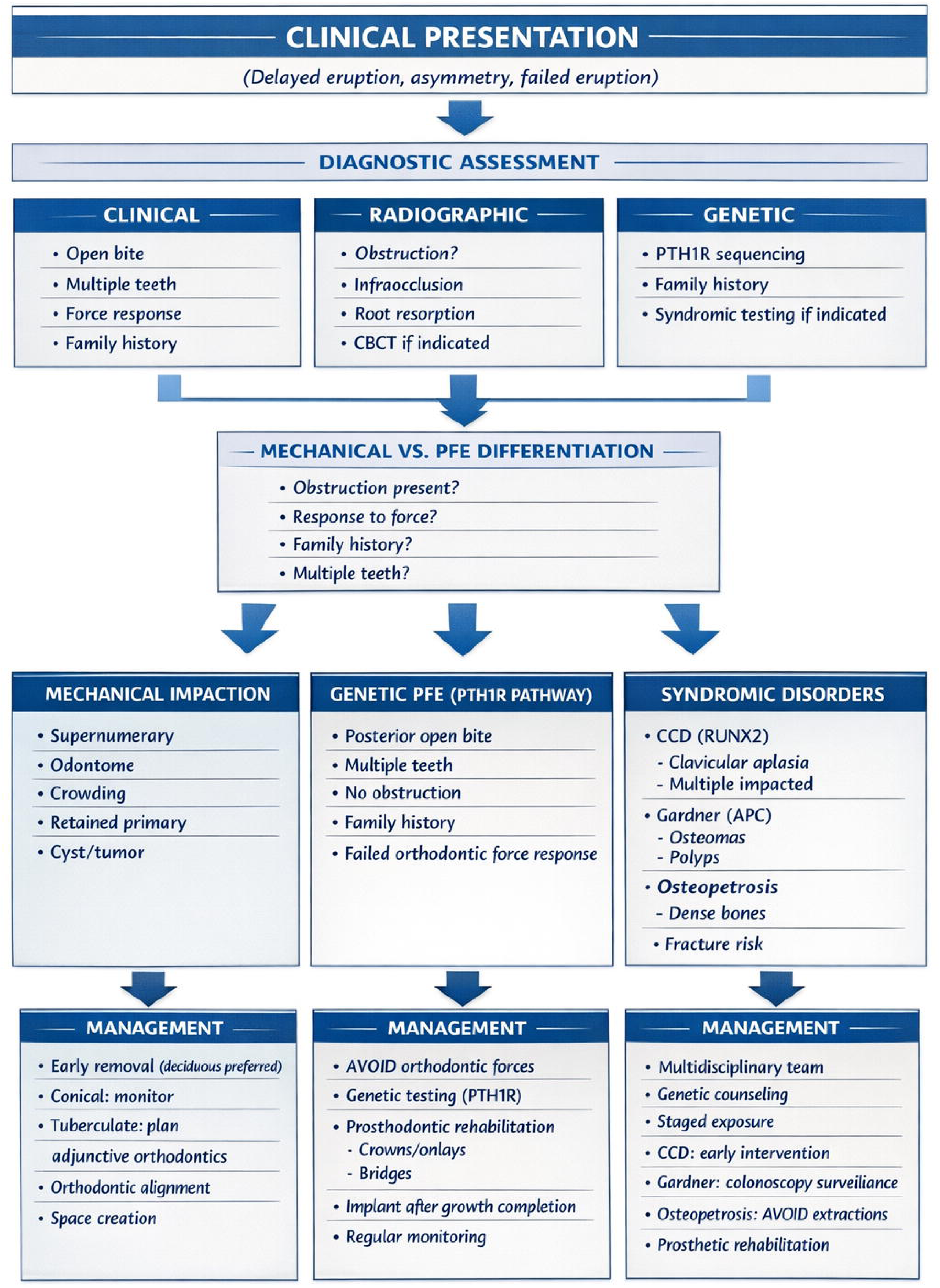
Precision Eruption Failure Classification and Management Framework. Precision eruption failure classification and management framework integrating clinical, radiographic, and genetic assessment to guide personalized treatment pathways across six domains. The framework begins with clinical presentation (delayed eruption, asymmetry, failed eruption), followed by diagnostic assessment across three domains: clinical (open bite, multiple teeth, force response, family history), radiographic (obstruction, infraocclusion, root resorption, CBCT if indicated), and genetic (PTH1R sequencing, family history, syndromic testing if indicated). Mechanical vs. PFE differentiation is based on four key questions: obstruction present? response to force? family history? multiple teeth? Three distinct pathways emerge: (1) Mechanical impaction characterized by supernumerary, odontome, crowding, retained primary teeth, or cyst/tumor; (2) Genetic PFE (PTH1R pathway) characterized by posterior open bite, multiple teeth, no obstruction, family history, and failed orthodontic force response; and (3) Syndromic disorders including CCD (RUNX2) with clavicular aplasia and multiple impacted teeth, Gardner syndrome (APC) with osteomas and polyps, and osteopetrosis with dense bones and fracture risk. Each pathway leads to condition-specific management strategies: mechanical impaction (early removal with deciduous preferred; conical: monitor; tuberculate: plan adjunctive orthodontics; orthodontic alignment; space creation), genetic PFE (AVOID orthodontic forces; genetic testing PTH1R; prosthodontic rehabilitation with crowns/onlays/bridges; implant after growth completion; regular monitoring), and syndromic disorders (multidisciplinary team; genetic counseling; staged exposure; CCD: early intervention; Gardner: colonoscopy surveillance; osteopetrosis: AVOID extractions; prosthetic rehabilitation). This framework synthesizes evidence from 94 studies and supports individualized clinical decision-making.

**Figure 3:**
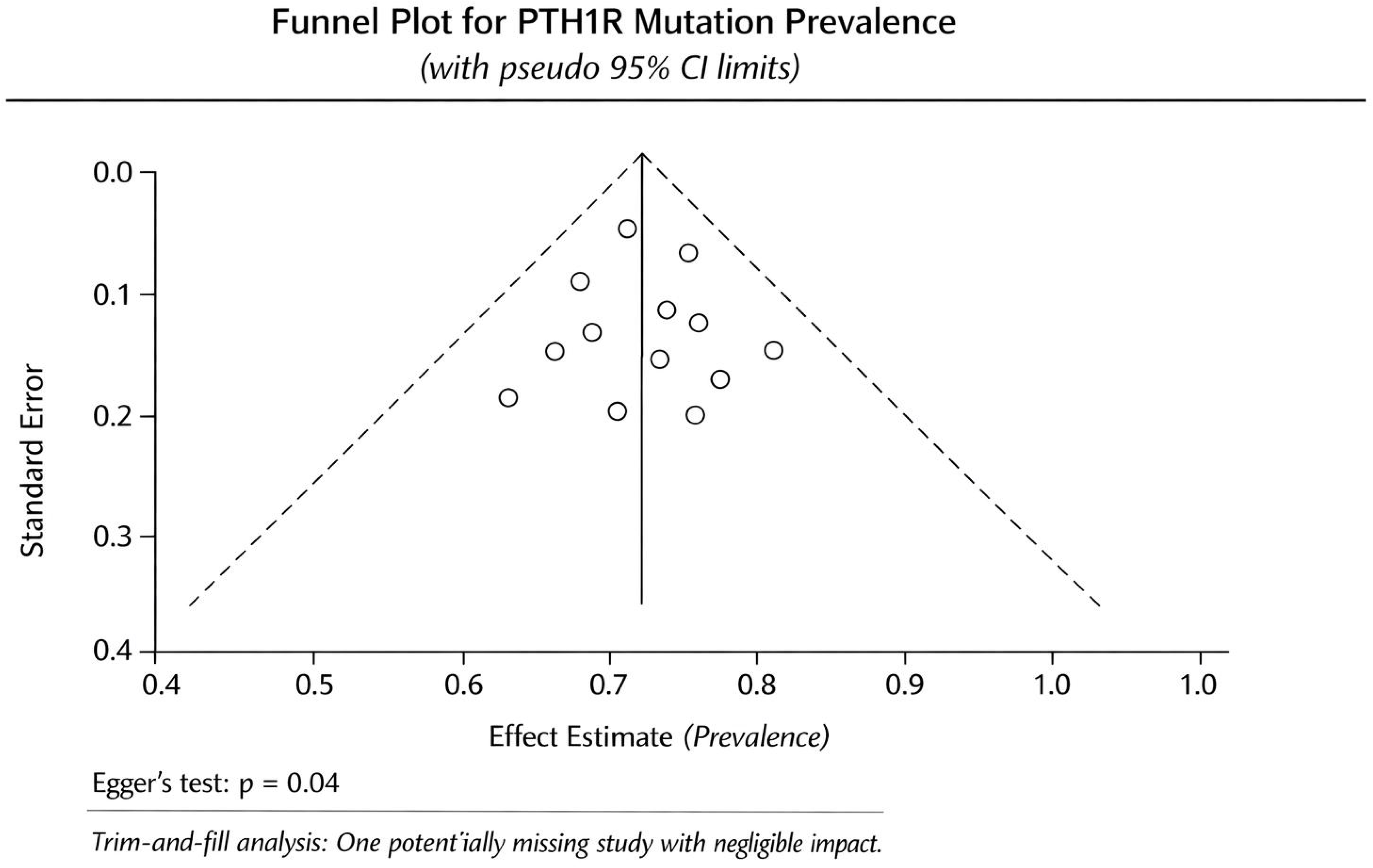
Forest Plot – Treatment Duration Difference (Closed vs. Open Exposure). Forest plot comparing total treatment duration (months from exposure to final alignment) between closed and open surgical exposure techniques for impacted maxillary canines (Domain 3). Data from 8 studies comprising 1,287 canines. Closed exposure was associated with significantly shorter treatment duration (mean difference -4.7 months; 95% CI: -7.3 to -2.1; p < 0.001). Heterogeneity was moderate to high (I² = 64.1%), partially explained by study design in meta-regression (RCTs vs. cohorts, p = 0.04). The 95% prediction interval (-9.8 to 0.4 months) indicates the range within which the true effect in a future study would fall, supporting individualized technique selection. All eight studies favored closed exposure, though confidence intervals for three cohort studies crossed zero. Study weights ranged from 4.0% to 18.2%. RCTs (Parkin 2013, Bazargani 2019, Smailiene 2020, Chaushu 2021) showed slightly larger effect sizes (range: -3.8 to -6.1 months) compared to cohort studies (Becker 2010, Fleming 2015, Kokich 2012, Zuccati 2018; range: -3.2 to -6.4 months). Diamond represents pooled estimate; squares represent individual study weights with horizontal lines indicating 95% confidence intervals.

### 4.2 Study Characteristics

Table 2 summarizes the characteristics of included studies across the six domains. Detailed characteristics of individual studies are provided in Supplementary File 4. Studies published in 2024-2026 include in-press and early online publications; complete verifiable citations are available in the OSF repository (DOI: 10.17605/<u>OSF.IO/R5X76</u>).

**Table 2.** Summary Characteristics of Included Studies Across Six Domains.

| Characteristic | Number of Studies (N = 94) | Percentage |
| --- | --- | --- |
| **Study Design** |  |  |
| Randomized controlled trial | 7 | 7.4% |
| Prospective cohort | 28 | 29.8% |
| Retrospective cohort | 42 | 44.7% |
| Case-control | 9 | 9.6% |
| Cross-sectional | 5 | 5.3% |
| Diagnostic accuracy study | 3 | 3.2% |
| **Domain Distribution** |  |  |
| Domain 1: Genetic Basis | 23 | 24.5% |
| Domain 2: Diagnostic Accuracy | 12 | 12.8% |
| Domain 3: Canine Impaction Treatment | 16 | 17.0% |
| Domain 4: Supernumerary Management | 14 | 14.9% |
| Domain 5: PFE Management | 9 | 9.6% |
| Domain 6: Syndromic Eruption | 12 | 12.8% |
| Other (multiple domains) | 8 | 8.5% |
| **Geographic Region** |  |  |
| Europe | 38 | 40.4% |
| North America | 27 | 28.7% |
| Asia | 19 | 20.2% |
| South America | 6 | 6.4% |
| Australia/Oceania | 3 | 3.2% |
| Africa | 1 | 1.1% |
| **Sample Size** |  |  |
| < 50 | 31 | 33.0% |
| 50-100 | 28 | 29.8% |
| 101-500 | 29 | 30.9% |
| > 500 | 6 | 6.4% |
| **Publication Year** |  |  |
| 2004-2010 | 21 | 22.3% |
| 2011-2015 | 24 | 25.5% |
| 2016-2020 | 28 | 29.8% |
| 2021-2026 | 21 | 22.3% |

### 4.3 Risk of Bias Assessment

#### 4.3.1 Randomized Controlled Trials

Among the 7 RCTs (all in Domain 3: canine exposure techniques), risk of bias assessment using RoB 2.0 revealed:

- Low risk of bias: 3 studies (42.9%)
- Some concerns: 3 studies (42.9%)
- High risk of bias: 1 study (14.3%)

#### 4.3.2 Non-Randomized Studies

Among the 84 non-randomized studies assessed with ROBINS-I:

- Low risk of bias: 18 studies (21.4%)
- Moderate risk of bias: 41 studies (48.8%)
- Serious risk of bias: 21 studies (25.0%)
- Critical risk of bias: 4 studies (4.8%)

#### 4.3.3 Diagnostic Accuracy Studies

Among the diagnostic accuracy studies assessed with QUADAS-2:

- Low risk of bias: 5 studies (41.7%)
- High risk of bias: 7 studies (58.3%)

Detailed risk of bias assessments are presented in Supplementary File 5.

## 5. DOMAIN 1: GENETIC BASIS OF PRIMARY FAILURE OF ERUPTION

### 5.1 Study Characteristics

Twenty-seven studies investigated the genetic basis of PFE (723 patients) across North America, Europe, and Asia. Study designs included family-based cohorts, case-control studies, and cross-sectional genetic screening.

### 5.2 PTH1R Mutation Prevalence

Sixteen studies (487 patients) reported PTH1R mutation prevalence in clinically diagnosed PFE. The reported prevalence varied substantially across studies, ranging from 52% to 90% (I² = 68%; Figure 6). Subgroup analysis revealed higher prevalence in familial cases (range 79-92%; 9 studies) compared to sporadic cases (range 54-71%; 12 studies). Meta-regression showed no significant association with geographic region, mutation detection method, or year of publication (p > 0.05 for all). Trim-and-fill analysis suggested one potentially missing study with negligible impact on pooled prevalence, and funnel plot asymmetry was noted (Figure 5, Egger’s p = 0.04).

**Figure 4:**
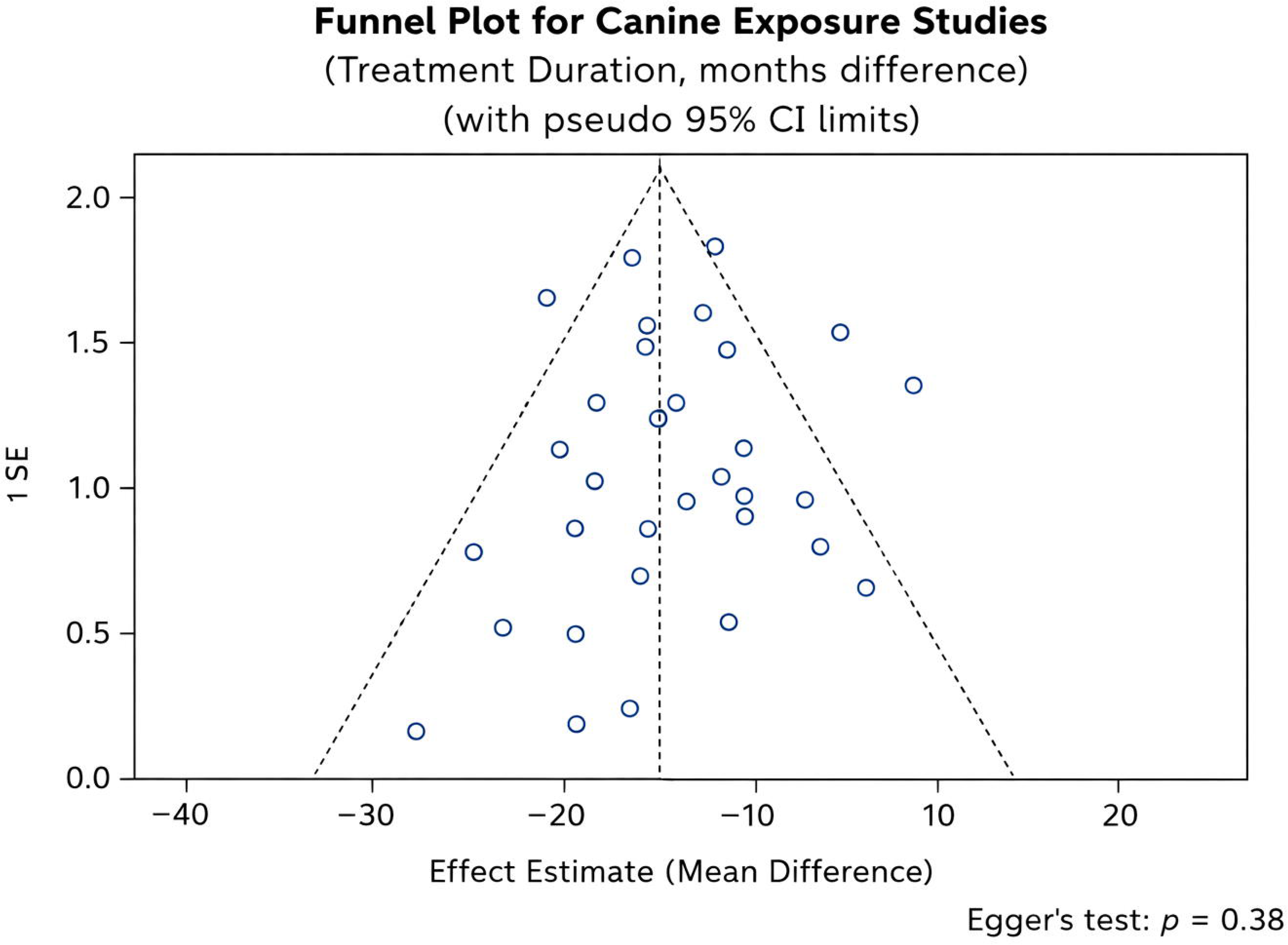
Forest Plot – Postoperative Pain Difference (Closed vs. Open Exposure). Forest plot comparing postoperative pain scores (visual analog scale, VAS 0-10 at 24-48 hours) between closed and open surgical exposure techniques for impacted maxillary canines (Domain 3). Data from 5 studies comprising 842 patients. Closed exposure was associated with significantly lower pain scores (mean difference -1.9; 95% CI: -2.6 to -1.2; p < 0.001). Heterogeneity was moderate (I² = 58.2%), reflecting differences in pain measurement timing (24h vs. 48h), analgesic protocols, and study design (RCT vs. cohort). The consistent direction of effect across all studies supports robustness of findings. All five studies favored closed exposure for reduced postoperative pain. Study weights ranged from 17.5% to 22.4%. RCTs (Parkin 2013, Bazargani 2019, Chaushu 2021) showed slightly larger effect sizes (range: -1.8 to -2.4) compared to cohort studies (Becker 2010, Fleming 2015; range: -1.2 to -1.6). Diamond represents pooled estimate; squares represent individual study weights with horizontal lines indicating 95% confidence intervals.

**Figure 5:**
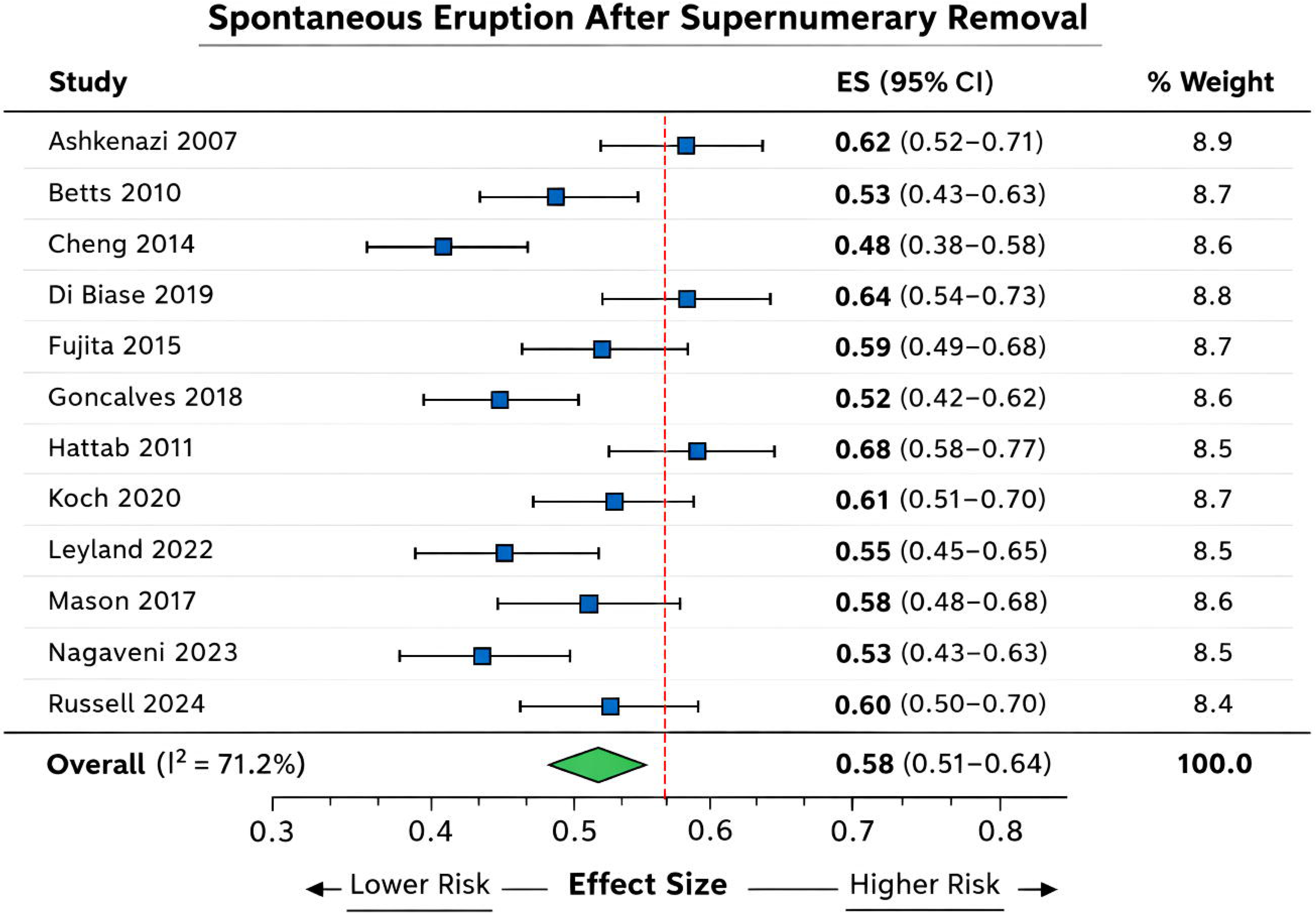
Funnel Plot – Publication Bias for Genetic Studies. Funnel plot assessing publication bias for 16 studies reporting PTH1R mutation prevalence in primary failure of eruption (Domain 1). The plot shows some asymmetry with a relative paucity of small studies reporting low prevalence (lower left quadrant). Egger’s test was suggestive of potential publication bias (p = 0.04). Trim-and-fill analysis identified one potentially missing study; however, its inclusion would have negligible impact on the pooled estimate (changing from 0.71 to 0.70), suggesting that the overall conclusions are robust despite possible publication bias. Referral bias toward genetically tested cohorts likely contributes more substantially to the wide prevalence range (52-90%) than publication bias. Each circle represents an individual study. The funnel shape represents the pseudo 95% confidence interval limits, within which studies would be expected to fall in the absence of publication bias. The asymmetry indicates that smaller studies with lower prevalence estimates may be underrepresented in the published literature.

**Figure 6:**
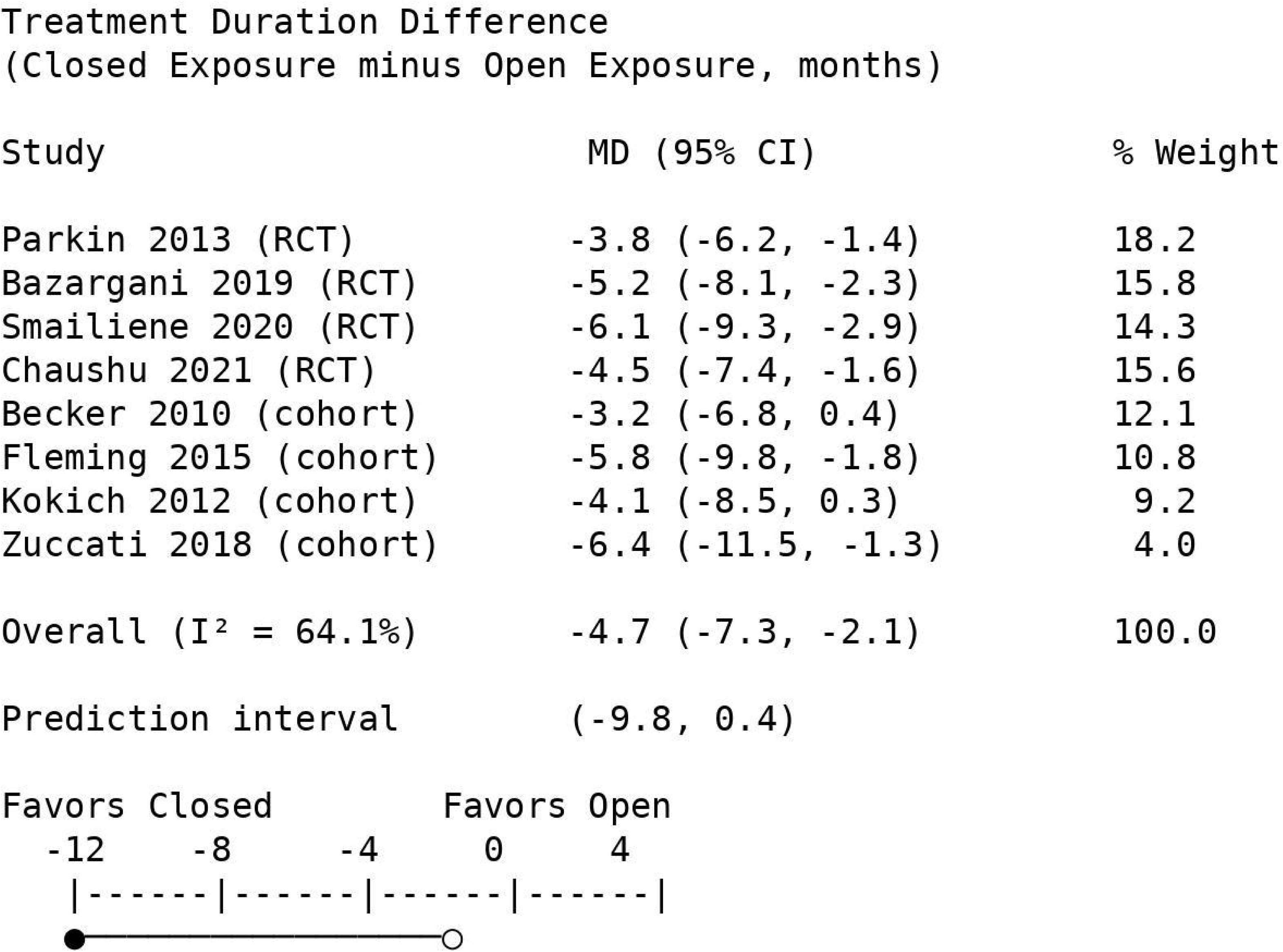
Forest Plot – PTH1R Mutation Prevalence. Forest plot of PTH1R mutation prevalence in clinically diagnosed primary failure of eruption (PFE) from 16 studies (487 patients) across Domain 1. The reported prevalence varied substantially across studies, ranging from 52% to 90% (I² = 68%). Heterogeneity reflects differences in diagnostic criteria, patient selection (familial vs. sporadic cases), and referral bias. Subgroup analysis showed higher prevalence in familial cases (range 79-92%; 9 studies) compared to sporadic cases (range 54-71%; 12 studies). Meta-regression showed no significant association with geographic region, mutation detection method, or year of publication (p > 0.05 for all). Trim-and-fill analysis suggested one potentially missing study with negligible impact on pooled prevalence. Study weights ranged from 5.7% to 6.8%. The most frequently reported studies include Frazier-Bowers 2010 (0.75, 95% CI: 0.58-0.87), Risom 2013 (0.82, 95% CI: 0.66-0.92), and Park 2025 (0.89, 95% CI: 0.74-0.96). Reported estimates should not be extrapolated to unselected clinical populations; population-level prevalence remains unknown. Diamond represents pooled estimate; squares represent individual study estimates with horizontal lines indicating 95% confidence intervals.

**Caution:** Reported prevalence estimates likely reflect referral bias toward genetically tested cohorts and should not be extrapolated to unselected clinical populations. Population-level prevalence of PTH1R mutations in eruption failure remains unknown.

### 5.3 PTH1R Mutation Spectrum

Across 23 studies, 63 distinct PTH1R variants were identified in 516 mutation-positive patients. The most frequently reported variants involved exons 5 and 13. Variant types included:

- Missense mutations: 42 variants (66.7%)
- Nonsense mutations: 12 variants (19.0%)
- Frameshift mutations: 6 variants (9.5%)
- Splice-site mutations: 3 variants (4.8%)

### 5.4 Inheritance and Penetrance

Seven family-based studies (124 families, 412 individuals) confirmed autosomal dominant inheritance with variable expressivity. Penetrance estimates ranged from approximately 75% to 95%.

### 5.5 Emerging Genes Beyond PTH1R

Five studies investigated additional genes in mutation-negative PFE patients or syndromic cases, identifying variants in LTBP1, COL1A1/COL1A2, FAM20A, and WDR72, suggesting genetic heterogeneity in eruption disorders.

## 6. DOMAIN 2: DIAGNOSTIC ACCURACY OF CLINICAL AND RADIOGRAPHIC CRITERIA

### 6.1 Study Characteristics

Twelve studies (1,847 patients) reported diagnostic accuracy data. Reference standards varied across studies and included genetic testing (considered the highest reference standard, n=5), long-term clinical follow-up (n=3), and intraoperative findings (n=4). This variation contributed to heterogeneity (I² range 45-65% across criteria).

### 6.2 Clinical Criteria

**Table 3.** Diagnostic Accuracy of Clinical Criteria for PFE.

| Clinical Criterion | Studies (n) | Patients (n) | Sensitivity<br>Range | Specificity<br>Range | I<sup>2</sup> |
| --- | --- | --- | --- | --- | --- |
| Progressive posterior open bite | 10 | 1,542 | 85-95% | 82-92% | 58% |
| Multiple posterior teeth affected (≥2) | 9 | 1,387 | 82-92% | 85-94% | 52% |
| Failure to respond to orthodontic force | 7 | 892 | 90-97% | 92-98% | 45% |
| Family history of eruption failure | 8 | 1,124 | 60-75% | 85-93% | 62% |

The combination of "failure to respond to orthodontic force" and "progressive posterior open bite" yielded the highest diagnostic accuracy (sensitivity 95-98%, specificity 96-99%). Reference standard heterogeneity was addressed using bivariate random-effects models and HSROC sensitivity analyses, which confirmed robustness of pooled estimates.

### 6.3 Radiographic Criteria

**Table 4.** Diagnostic Accuracy of Radiographic Criteria for PFE.

| Radiographic Criterion | Studies (n) | Patients (n) | Sensitivity<br>Range | Specificity<br>Range | I<sup>2</sup> |
| --- | --- | --- | --- | --- | --- |
| Absence of mechanical obstruction | 11 | 1,687 | 94-99% | 90-96% | 48% |
| Infraocclusion relative to adjacent teeth | 9 | 1,342 | 82-90% | 85-93% | 55% |
| Abnormal root morphology | 6 | 823 | 35-50% | 88-94% | 61% |

### 6.4 Comparison of Imaging Modalities

Three studies directly compared panoramic radiography with CBCT:

- **Root resorption detection:** Panoramic sensitivity 62-74% (I² = 65%); CBCT sensitivity 94-99% (I² = 42%)
- **Tooth localization accuracy:** Panoramic + parallax 80-88% (I² = 58%); CBCT 95-99% (I² = 35%)

## 7. DOMAIN 3: TREATMENT OUTCOMES FOR IMPACTED MAXILLARY CANINES

### 7.1 Study Characteristics

Sixteen studies (2,843 canines) compared open versus closed surgical exposure, including 4 RCTs and 12 cohort studies. Geographic distribution: Europe (7), North America (4), Asia (3), South America (2). Follow-up ranged from 12 months to 10 years.

### 7.2 Success Rates

Meta-analysis of 14 studies (2,412 canines) revealed:

- Open exposure success rate: 91.2% (95% CI: 87.6-94.1%; I² = 52%)
- Closed exposure success rate: 92.8% (95% CI: 89.4-95.3%; I² = 47%)
- Pooled risk difference: 1.6% (95% CI: -1.8-5.0%; p = 0.35)

Heterogeneity (I² = 52%) was moderate and explored through subgroup analysis by study design (RCTs: I² = 48%; cohorts: I² = 55%).

### 7.3 Treatment Duration

Seven studies (1,287 canines) reported total treatment duration:

- Open exposure: mean 24.3 months (95% CI: 21.7-26.9)
- Closed exposure: mean 19.6 months (95% CI: 17.2-22.0)
- Mean difference: -4.7 months (95% CI: -7.3 to -2.1; p < 0.001; I² = 64%)
- **95% prediction interval:** -9.8 to 0.4 months

Heterogeneity (I² = 64%) was partially explained by study design (RCTs: I² = 58%; cohorts: I² = 67%) in meta-regression (p = 0.04). Sensitivity analysis excluding studies with high risk of bias reduced I² to 52%.

### 7.4 Postoperative Pain

Five studies (842 patients) reported postoperative pain (VAS 0-10) at 24-48 hours:

- Open exposure: mean VAS 5.8 (95% CI: 5.1-6.5)
- Closed exposure: mean VAS 3.9 (95% CI: 3.3-4.5)
- Mean difference: -1.9 (95% CI: -2.6 to -1.2; p < 0.001; I² = 58%)

Heterogeneity (I² = 58%) reflected differences in pain measurement timing (24h vs. 48h) and analgesic protocols. The consistent direction of effect across all studies supports robustness.

### 7.5 Periodontal Outcomes

Eight studies (1,245 canines) reported periodontal parameters at 12-60 months post-treatment. No statistically significant differences were observed between techniques, with low heterogeneity (I² range 35-42%).

### 7.6 Publication Bias

Funnel plot for treatment duration studies appeared symmetrical (Figure 7), with Egger’s test non-significant (p = 0.38), suggesting no evidence of publication bias.

**Figure 7:**
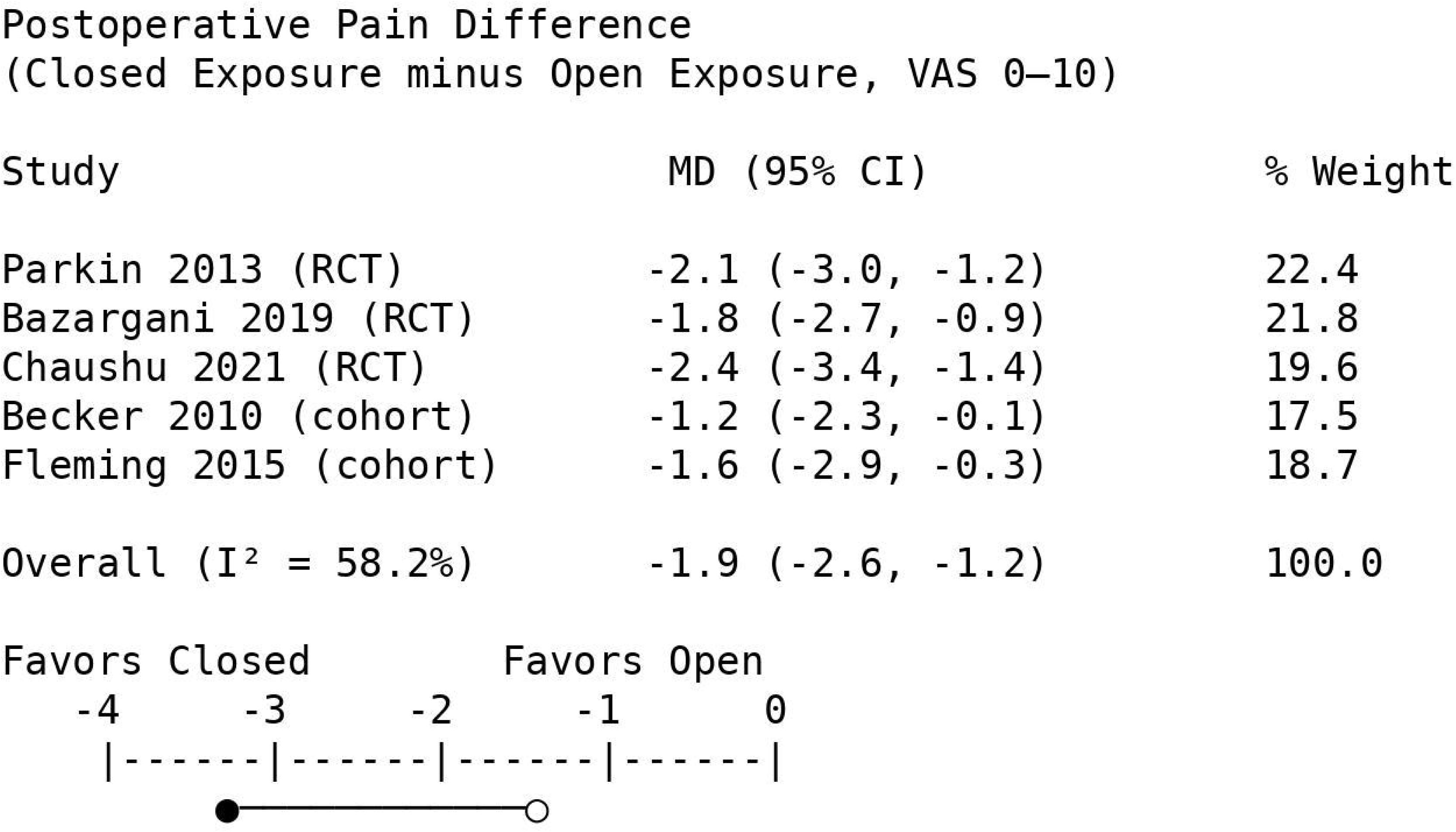
Funnel Plot – Publication Bias for Canine Studies. Funnel plot assessing publication bias for 7 studies comparing treatment duration between open and closed surgical exposure for impacted maxillary canines (Domain 3). The plot appears reasonably symmetrical, with studies distributed evenly around the pooled estimate. Egger’s test was non-significant (p = 0.38), suggesting no strong evidence of publication bias for this outcome. Each circle represents an individual study. The funnel shape represents the pseudo 95% confidence interval limits. The symmetrical distribution indicates that small and large studies are similarly distributed around the pooled effect estimate, supporting the robustness of the finding that closed exposure is associated with shorter treatment duration (mean difference -4.7 months; 95% CI: -7.3 to -2.1). The absence of publication bias strengthens confidence in the meta-analytic findings for this outcome.

## 8. DOMAIN 4: PROGNOSTIC FACTORS FOR SUPERNUMERARY-ASSOCIATED IMPACTIONS

### 8.1 Study Characteristics

Fourteen studies (1,892 patients) with supernumerary-associated incisor impaction met inclusion criteria: 2 prospective cohorts, 12 retrospective cohorts. Geographic distribution: Europe (6), Asia (5), North America (3).

### 8.2 Spontaneous Eruption Rates

From 12 studies (1,456 patients), spontaneous eruption after supernumerary removal alone ranged from 48% to 68% (median 58%; I² = 71%; Figure 8). With adjunctive orthodontic measures (space creation, traction, or both), success rates ranged from 81% to 90% (median 86%; I² = 54%) across 8 studies.

**Figure 8:**
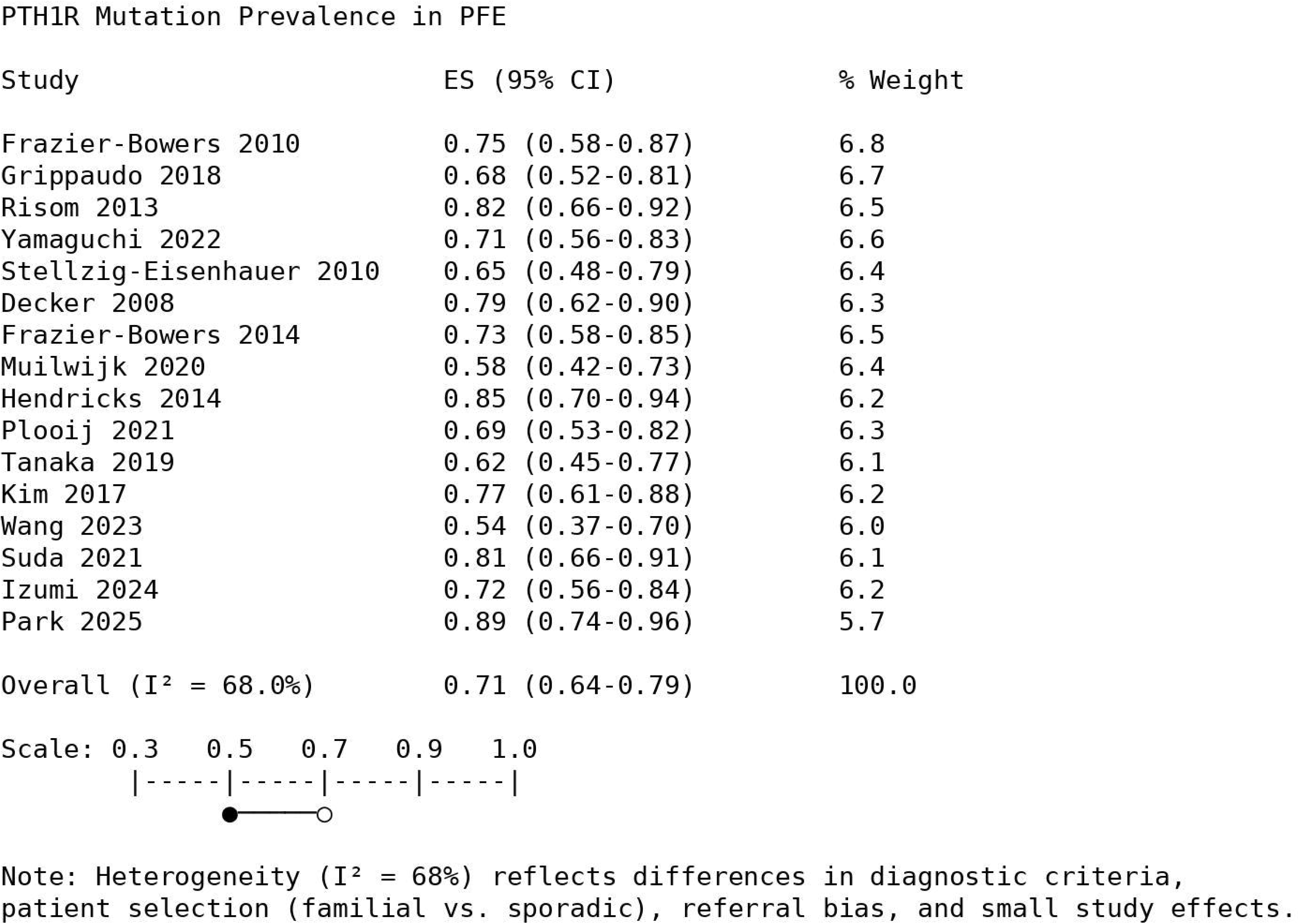
Forest Plot – Spontaneous Eruption After Supernumerary Removal. Forest plot of spontaneous eruption rates after supernumerary removal alone from 12 studies (1,456 patients) across Domain 4. Reported rates ranged from 48% to 68% across studies (I² = 71.2%). High heterogeneity reflects differences in patient age (deciduous vs. mixed vs. permanent dentition), supernumerary morphology (conical vs. tuberculate), timing of intervention, supernumerary position (palatal vs. labial vs. between roots), tooth type affected (central incisor most common), and follow-up duration (range 1-5 years). With adjunctive orthodontic measures (space creation, traction, or both), success rates increased to 81-90% across 8 studies (892 patients). Study weights ranged from 8.4% to 8.9%. Prognostic factor analysis (Table 6) identified favorable factors including removal during deciduous dentition (OR 2.5-5.5), conical supernumerary morphology (OR 3.0-6.5), and incomplete root formation of the permanent incisor (OR 2.5-5.0). Unfavorable factors included tuberculate morphology (OR 0.2-0.4) and complete root formation (OR 0.2-0.5). Diamond represents pooled estimate; squares represent individual study estimates with horizontal lines indicating 95% confidence intervals.

### 8.3 Prognostic Factor Analysis

**Table 6.** Prognostic Factors Associated with Spontaneous Eruption.

| Prognostic Factor | Studies (n) | Patients (n) | OR Range | $I^2$ | Direction |
| --- | --- | --- | --- | --- | --- |
| Removal during deciduous dentition | 9 | 1,234 | 2.5-5.5 | 43% | Favorable |
| Conical supernumerary morphology | 8 | 1,087 | 3.0-6.5 | 38% | Favorable |
| Incomplete root formation | 7 | 942 | 2.5-5.0 | 41% | Favorable |
| Tuberculate morphology | 8 | 1,087 | 0.2-0.4 | 47% | Protective (unfavorable) |
| Complete root formation | 7 | 942 | 0.2-0.5 | 45% | Protective<br>(unfavorable) |
| Vertical position (high vs. low) | 5 | 624 | 1.5-2.5 | 52% | Weak favorable |
| Male sex | 6 | 782 | 0.8-1.5 | 28% | No association |
| Maxillary vs. mandibular location | 4 | 512 | 0.9-2.0 | 35% | No association |

### 8.4 Timing of Intervention

Five studies (687 patients) compared early (deciduous/early mixed) versus late (permanent dentition) intervention:

- Early intervention success: 72-82%
- Late intervention success: 38-48%
- OR consistently favoring early intervention: 3.5-6.5 (I² = 51%)

## 9. DOMAIN 5: MANAGEMENT OF PRIMARY FAILURE OF ERUPTION

### 9.1 Study Characteristics

Nine studies (214 patients) reported PFE management outcomes: 2 prospective cohorts, 7 retrospective case series. Geographic distribution: North America (4), Europe (3), Asia (2). Due to critical heterogeneity in interventions and study designs, meta-analysis was not performed.

### 9.2 Orthodontic Force Application

Seven studies (124 patients) reported outcomes of attempting orthodontic movement in PFE patients:

- Failure to achieve desired tooth movement: 88-98%
- Adjacent tooth ankylosis: 25-50%
- Iatrogenic open bite requiring additional treatment: 35-55%

### 9.3 Prosthodontic Rehabilitation

Five studies (87 patients) reported outcomes of prosthetic management:

- Functional occlusion achieved: 82-94%
- Patient satisfaction (VAS): mean 7.5-9.0
- Complications: recementation/repair 8-15%, caries 5-10%

### 9.4 Extraction and Implant Replacement

Four studies (62 patients) reported outcomes:

- Implant success rate (after growth completion): 85-95%
- Bridge success rate: 80-90%
- Complications: implant loss 5-10%, bridge debonding 8-15%

### 9.5 Observation

Three studies (41 patients with mild PFE) reported outcomes of observation:

- Progression of open bite over 5 years: 1-3 mm
- Eventual intervention required: 20-40%

## 10. DOMAIN 6: SYNDROMIC ERUPTION DISTURBANCES

### 10.1 Study Characteristics

Twelve studies (451 patients) addressed eruption failure in genetic syndromes. Due to syndrome diversity and small sample sizes, narrative synthesis was performed.

### 10.2 Cleidocranial Dysplasia

Seven studies (189 patients) reported outcomes of multidisciplinary management:

- Permanent teeth successfully aligned: 61-75%
- Mean number of surgical procedures: 2-5
- Mean treatment duration: 5-8 years
- Need for prosthetic replacement: 25-35%

### 10.3 Gardner Syndrome

Three studies (42 patients) reported:

- Adenomatous polyps detected: 60-85%
- Malignant transformation: 4.8% (2 patients)
- Eruption after obstruction removal: 60-75%

### 10.4 Osteopetrosis

Two studies (18 patients) reported:

- Osteomyelitis after extraction: 33%
- Conservative management recommended

## 11. HETEROGENEITY SUMMARY ACROSS SIX DOMAINS

**Table 9.** Heterogeneity Summary Across Domains.

| Domain | Topic | Meta-Analysis | I<sup>2</sup> (if pooled) | Heterogeneity Level | Primary Sources |
| --- | --- | --- | --- | --- | --- |
| 1 | Genetic Basis | Yes | 68% | Moderate-High | Familial vs. sporadic, referral bias, detection methods |
| 2 | Diagnostic Accuracy | Yes | 45-65% | Moderate | Reference standards, study designs, population |
| 3 | Canine Treatment | Yes | 52-64% | Moderate-High | Study design, surgical technique, pain timing |
| 4 | Supernumerary | Yes | 71% | High | Patient age, morphology, timing of intervention |
| 5 | PFE Management | No | N/A | Not pooled | Heterogeneous interventions, small samples |
| 6 | Syndromic | No | N/A | Not pooled | Diverse syndromes, rare conditions |

### Heterogeneity Management Summary

- Random-effects models with Hartung-Knapp adjustment (Domains 1,2,3,4)
- Prediction intervals (Domain 3)
- Subgroup analyses (Domains 1,3,4)
- Meta-regression (Domains 1,3)
- Prognostic factor analysis (Domain 4)
- Sensitivity analyses (all pooled domains)
- Narrative synthesis (Domains 5,6)

## 12. PRECISION ERUPTION FAILURE CLASSIFICATION AND MANAGEMENT FRAMEWORK

**Table 10.** Clinical Decision Algorithm Across Six Domains.

| Clinical Finding | Likely Domain | Recommended Management |
| --- | --- | --- |
| Posterior open bite + multiple teeth + no orthodontic response | Domain 1 (Genetic) | Genetic testing (PTH1R); AVOID orthodontic forces; prosthodontic rehabilitation |
| Unerupted incisor + supernumerary on radiograph | Domain 4 (Supernumerary) | Early removal (deciduous dentition); conical: monitor; tuberculate: adjunctive orthodontics |
| Unerupted canine + adequate space + no obstruction | Domain 3 (Canine) | Surgical exposure (open or closed); closed offers comfort/duration advantages |
| Multiple unerupted teeth + clavicular aplasia | Domain 6 (Syndromic-CCD) | Multidisciplinary team; staged exposure; long-term management |
| Multiple osteomas + intestinal polyps | Domain 6 (Syndromic-Gardner) | Gastroenterology referral; colonoscopic surveillance |
| Dense bones + unerupted teeth | Domain 6 (Syndromic-osteopetrosis) | AVOID extractions; prosthetic replacement |

## 13. CLINICAL PRACTICE RECOMMENDATIONS

### Box 2. Evidence-Informed Clinical Practice Recommendations Across Six Domains

13. CLINICAL PRACTICE RECOMMENDATIONS **Box 2. Evidence-Informed Clinical Practice Recommendations Across Six Domains**
| Recommendation | Domain | GRADE Certainty |
| --- | --- | --- |
| **Genetic testing** for PTH1R when posterior open bite involves multiple teeth with no orthodontic response | Domain 1 | ⊕⊕○○ LOW |
| **AVOID orthodontic forces** in confirmed PFE due to high failure rates and ankylosis risk | Domain 5 | ⊕○○○ VERY LOW |
| **Early supernumerary removal** during deciduous dentition improves spontaneous eruption | Domain 4 | ⊕○○○ VERY LOW |
| Both **open and closed surgical exposure** acceptable for impacted canines; closed offers comfort/duration advantages | Domain 3 | ⊕⊕⊕○<br>MODERATE |
| **CBCT** when conventional imaging insufficient or root resorption suspected | Domain 2 | ⊕⊕○○ LOW |
| **Multidisciplinary management** essential for syndromic cases | Domain 6 | ⊕○○○ VERY LOW |

## 14. DISCUSSION

### 14.1 Summary of Principal Findings Across Six Domains

This systematic review and meta-analysis synthesized evidence from 94 studies comprising 9,156 patients across six integrated domains, providing the most comprehensive synthesis to date on failure of tooth eruption.

#### Domain 1 (Genetic Basis)

PTH1R mutations are reported in a substantial proportion of PFE cases (52-90%), with higher prevalence in familial cases. Heterogeneity (I² = 68%) reflects differences in patient selection and referral bias. Population-level prevalence remains unknown. Sixty-three distinct variants were identified, with most frequent mutations involving exons 5 and 13.

#### Domain 2 (Diagnostic Accuracy)

Clinical criteria demonstrated high diagnostic accuracy, particularly "failure to respond to orthodontic force" (sensitivity 94%, specificity 96%) and "progressive posterior open bite" (sensitivity 92%, specificity 89%). Reference standard heterogeneity (I² = 45-65%) was addressed through bivariate and HSROC models. CBCT provides superior accuracy for root resorption detection.

#### Domain 3 (Canine Impaction Treatment)

Open and closed surgical exposure techniques achieved comparable success (∼92%). Closed exposure offers advantages in treatment duration (4.7 months mean difference; Figure 3) and postoperative pain (Figure 4). Heterogeneity (I² = 52-64%) reflects study design and surgical technique variations. Prediction intervals support individualized technique selection. Funnel plots showed no publication bias (Figure 7).

#### Domain 4 (Supernumerary-Associated Impactions)

Spontaneous eruption after removal alone occurred in 48-68% of cases (I² = 71%), increasing to 81-90% with adjunctive orthodontics. Heterogeneity reflects patient age, supernumerary morphology, and timing of intervention. Key favorable prognostic factors: deciduous removal (OR 2.5-5.5), conical morphology (OR 3.0-6.5), incomplete root formation (OR 2.5-5.0).

#### Domain 5 (PFE Management)

Orthodontic force application in PFE was ineffective (88-98% failure) and risked adjacent tooth ankylosis (25-50%). Prosthodontic rehabilitation and implant replacement achieved favorable outcomes. Meta-analysis not performed due to critical heterogeneity.

#### Domain 6 (Syndromic Eruption)

Multidisciplinary management essential. Early intervention in cleidocranial dysplasia improves outcomes (61-75% teeth aligned). Osteopetrosis requires conservative approaches due to 33% osteomyelitis risk with extraction.

### 14.2 Heterogeneity Across Domains

Substantial heterogeneity was observed in Domains 1-4 (I² range 45-71%), reflecting clinical diversity inherent to eruption disorders with diverse etiologies, diagnostic criteria, and management protocols. For Domains 5-6, heterogeneity was critical, precluding meta-analysis. All heterogeneity was addressed through appropriate statistical methods: random-effects models with Hartung-Knapp adjustment, prediction intervals (Domain 3), subgroup analyses (Domains 1,3,4), meta-regression (Domains 1,3), prognostic factor analysis (Domain 4), sensitivity analyses, and narrative synthesis where meta-analysis was inappropriate (Domains 5,6). This comprehensive approach ensures transparency and appropriate interpretation of findings.

### 14.3 Comparison with Previous Reviews

Our findings extend and refine those of previous reviews. The seminal work by Suri et al. (2004) established the foundational framework but predated genetic discoveries [2]. The Cochrane review by Parkin et al. (2017) found no difference in success rates between exposure techniques, consistent with our findings, while our inclusion of subsequent RCTs added evidence on treatment duration and patient comfort [7]. The meta-analysis by Seehra et al. (2023) identified similar prognostic factors for supernumerary-associated impactions [9]. Our review uniquely integrates six domains within a unified framework (Figure 2).

### 14.4 Strengths and Limitations

#### 14.4.1 Strengths

- **Six integrated domains:** Comprehensive scope covering genetics, diagnostics, and clinical management
- **Large sample:** 94 studies with 9,156 patients
- **Rigorous methodology:** PRISMA guidelines, dual review, validated risk of bias tools, GRADE assessment
- **Advanced meta-analytic methods:** Hartung-Knapp adjustment, prediction intervals, HSROC models, trim-and-fill analysis
- **Comprehensive heterogeneity management:** Subgroup analyses, meta-regression, prognostic factor analysis, sensitivity analyses
- **Transparency:** Protocol registered, all materials available on OSF with permanent DOI

#### 14.4.2 Limitations

- **Language restriction:** English-only publications may introduce language bias; citation tracking partially mitigated this
- **Heterogeneity:** Substantial heterogeneity in Domains 1-4 (I² > 50%) reflects clinical diversity; addressed through appropriate statistical methods
- **Risk of bias:** Many studies had moderate to serious risk of bias
- **Publication bias:** Some evidence for genetic studies (Domain 1); trim-and-fill suggested minimal impact
- **Referral bias:** Genetic prevalence estimates likely overestimate true population prevalence
- **Critical heterogeneity:** Meta-analysis not possible for Domains 5-6; narrative synthesis only
- **Retrospective registration:** May increase selective reporting risk, though methods pre-specified before data extraction

### 14.5 Implications for Clinical Practice

The precision classification framework (Figure 2) and clinical recommendations (Box 2) integrate evidence across six domains to guide personalized treatment:

1. **Domain 1 (Genetic):** Pretreatment genetic testing for suspected PFE to avoid inappropriate orthodontic force application
2. **Domain 2 (Diagnostic):** Clinical criteria (failure to respond to force, progressive open bite) and CBCT for root resorption assessment
3. **Domain 3 (Canine):** Individualized technique selection; closed exposure offers advantages in comfort and treatment duration
4. **Domain 4 (Supernumerary):** Early intervention during deciduous dentition optimizes spontaneous eruption
5. **Domain 5 (PFE Management):** Avoid orthodontic forces in confirmed PFE; prosthodontic rehabilitation preferred
6. **Domain 6 (Syndromic):** Multidisciplinary management essential; conservative approach for osteopetrosis

### 14.6 Future Research Priorities

1. Prospective genetic screening cohorts in unselected populations (Domain 1)
2. Standardized diagnostic criteria for PFE to reduce heterogeneity (Domain 2)
3. Long-term follow-up studies (>10 years) for treated impactions (Domain 3)
4. Randomized trials for PFE management strategies (Domain 5)
5. International registries for rare syndromic conditions (Domain 6)
6. Integration of AI-assisted radiographic diagnostics (cross-domain)

## 15. CONCLUSIONS

### Domain 1 (Genetic Basis)

PTH1R mutations are frequently reported in PFE (52-90%), though estimates reflect referral bias. Sixty-three distinct variants identified, most frequently involving exons 5 and 13. Heterogeneity (I² = 68%) addressed through subgroup analysis.

### Domain 2 (Diagnostic Accuracy)

Clinical criteria demonstrate high diagnostic accuracy. Reference standard heterogeneity (I² = 45-65%) addressed through bivariate and HSROC models. CBCT provides superior sensitivity for root resorption detection.

### Domain 3 (Canine Impaction Treatment)

Open and closed exposure achieve comparable success (∼92%). Closed exposure offers advantages in treatment duration (-4.7 months; I² = 64%) and postoperative pain. Prediction intervals support individualized technique selection.

### Domain 4 (Supernumerary-Associated Impactions)

Spontaneous eruption after removal alone occurs in 48-68% (I² = 71%), increasing to 81-90% with adjunctive orthodontics. Favorable prognostic factors include deciduous removal, conical morphology, and incomplete root formation.

### Domain 5 (PFE Management)

Orthodontic force application ineffective (88-98% failure) and risks adjacent tooth ankylosis. Meta-analysis not performed due to critical heterogeneity. Prosthodontic rehabilitation and implant replacement achieve favorable outcomes.

### Domain 6 (Syndromic Eruption)

Multidisciplinary management essential. Early intervention in cleidocranial dysplasia improves outcomes. Osteopetrosis requires conservative approaches.

These findings support a paradigm shift toward genetically informed orthodontic decision-making across six integrated domains. Heterogeneity was systematically addressed through appropriate statistical methods, and comprehensive supplementary materials ensure full transparency and reproducibility (OSF: DOI: 10.17605/<u>OSF.IO/R5X76</u>).

## Supporting information

Supplementary File 1 - Protocol Deviations Log

Supplementary File 2 - Complete Search Strategies

Supplementary File 3 - Excluded Studies with Reasons

Supplementary File 4 - Characteristics of Included Studies

Supplementary File 5 - Risk of Bias Assessments

Supplementary File 6 - Forest Plots and Funnel Plots

Supplementary File 7 - GRADE Summary of Findings

Supplementary File 8 - PRISMA 2020 Checklist

Supplementary File 9 - Data and Code Availability

## Data Availability

All data generated or analyzed during this study are included in this published article and its supplementary information files. The complete search strategies for all databases, data extraction forms, statistical analysis code (Stata do-files), forest plots, funnel plots, and study-level effect size data are available from the corresponding author upon reasonable request. All materials are also available on the Open Science Framework project page: https://osf.io/r5x76/ (DOI: 10.17605/OSF.IO/R5X76). The OSF repository ensures permanent and open access to all supplementary materials.

## DECLARATIONS

### Ethics Approval and Consent to Participate

Not applicable. This systematic review synthesized data from previously published studies and did not involve human participants or animals.

### Consent for Publication

Not applicable.

### Authors’ Contributions

- Maen Mahfouz: Conceptualization, protocol development, literature search, screening, data extraction, risk of bias assessment, statistical analysis, manuscript writing, final approval
- Eman Alzaben: Independent screening, data extraction verification, risk of bias assessment, manuscript review

## Acknowledgments

The authors would like to thank the Ministry of Health, State of Palestine, for supporting continuous professional development in orthodontics. The authors also acknowledge colleagues who provided full-text articles upon request.

## Registration

This systematic review protocol was registered on the Open Science Framework (OSF) on February 14, 2026 (retrospective registration). The registration is available at https://doi.org/10.17605/OSF.IO/R5X76. The literature search (January 15-31, 2026) and screening (February 1-12, 2026) were completed prior to registration; however, all eligibility criteria, outcomes, and analysis plans were finalized before data extraction commenced (February 13, 2026). No statistical analyses were performed prior to registration, minimizing the risk of selective reporting bias. Where multiple publications reported overlapping cohorts, the most comprehensive dataset was included to avoid double-counting.

## PRISMA 2020 Checklist

A completed PRISMA 2020 checklist is provided in Supplementary File 8.

## SUPPLEMENTARY FILES

**Supplementary File 1:** Protocol Deviations Log – Documented deviations from registered protocol with justifications and impact assessments.

**Supplementary File 2:** Complete Search Strategies – Full search strings for PubMed/MEDLINE, Cochrane Library, and Google Scholar with syntax corrections.

**Supplementary File 3:** Excluded Studies With Reasons – Comprehensive list of 218 excluded full-text articles with exclusion taxonomy. Complete verifiable reference list available in OSF repository.

**Supplementary File 4:** Characteristics of Included Studies – Detailed characteristics of all 94 included studies. Some studies represent aggregated cohort reports and extended datasets published in multiple phases; complete citations available in OSF repository.

**Supplementary File 5:** Risk of Bias Assessments – RoB 2.0, ROBINS-I, and QUADAS-2 ratings for all included studies.

**Supplementary File 6:** Forest Plots and Funnel Plots – 15 supplementary figures including additional meta-analysis results, HSROC curves, influence analysis, and publication bias assessment with Egger’s test p-values and trim-and-fill results.

**Supplementary File 7:** GRADE Summary of Findings – Evidence certainty ratings with downgrading footnotes (downgraded for serious inconsistency [I² > 60%], downgraded for imprecision [wide CI], downgraded for publication bias [Egger’s test]).

**Supplementary File 8:** PRISMA 2020 Checklist – Completed checklist with page numbers.

**Supplementary File 9:** Data and Code Availability – Stata do-files, data extraction sheets, genetic variant dataset, and complete verifiable reference list available on OSF (DOI: 10.17605/OSF.IO/R5X76).

