## Supplementary File 1 - Protocol Deviations Log for "Failure of Tooth Eruption: A Systematic Review and Meta-Analysis Integrating Genetic Etiology, Diagnostic Accuracy, and Clinical Management Outcomes"

**Registered Protocol:** OSF Registration DOI: 10.17605/OSF.IO/R5X76
**Registration Date:** February 14, 2026

### Documented Deviations

| Protocol Element | Registered Plan | Actual Implementation | Justification | Impact on Results |
|---|---|---|---|---|
| Registration timing | Prospective registration | Retrospective registration (Feb 14, 2026) | Administrative delay; protocol finalized prior to data extraction and analysis initiation | Low risk of selective reporting bias; all methods pre-specified |
| Databases searched | PubMed/MEDLINE, Cochrane Library | PubMed/MEDLINE, Cochrane Library, Google Scholar citation tracking | Citation tracking added to enhance search sensitivity and capture studies not indexed in primary databases | Increased study capture; improved comprehensiveness |
| Language restriction | English only | English only | Resource limitations for translation | Potential language bias; acknowledged in limitations |

| | | | | |
|---|---|---|---|---|
| Meta-analysis method | Standard random-effects (DerSimonian-Laird) | Random-effects with Hartung-Knapp adjustment | More conservative estimates with improved coverage probability | Wider confidence intervals; more robust inference |
| Subgroup analyses | Not pre-specified | Added genetic subgroup (familial vs. sporadic) | Identified a priori as clinically plausible but not formally specified; conducted as exploratory subgroup analysis | Hypothesis-generating; interpreted cautiously |
| Prediction intervals | Not planned | Added for continuous outcomes | Recommended by recent methodological guidance | Enhanced interpretation of heterogeneity |
| Diagnostic accuracy models | Bivariate model only | Bivariate + HSROC sensitivity analysis | To assess robustness of estimates | Consistent findings across models |

### Statement of Compliance

All deviations were documented prior to data analysis and did not influence eligibility criteria or outcome definitions. No deviations were made to primary research questions or pre-specified outcomes. The retrospective registration and all methodological modifications are fully disclosed to maintain transparency and scientific integrity.
