## Supplementary File 2 - Complete Search Strategies for "Failure of Tooth Eruption: A Systematic Review and Meta-Analysis Integrating Genetic Etiology, Diagnostic Accuracy, and Clinical Management Outcomes"

### PubMed/MEDLINE (Search Date: February 10, 2026)

**Interface:** National Library of Medicine (pubmed.ncbi.nlm.nih.gov)
**Results:** 1,892 records
**Coverage:** January 2004 – February 2026

#1 "Tooth Eruption"[MeSH Terms] OR "Tooth Eruption"[Title/Abstract] OR "Tooth Eruption, Ectopic"[MeSH Terms] OR "Ectopic Tooth Eruption"[Title/Abstract]

**Interface:** scholar.google.com
**Results:** 1,450 records screened (first 200 per structured search + citation tracking)
**Method:** Used only for supplementary citation tracking; screening restricted to first 200 results per query to minimize algorithmic bias and ensure feasibility, consistent with methodological guidance [13].

#### Structured Search Strings (800 records)

| Search Query | Results Screened | Rationale |
|---|---|---|

| | | |
|---|---|---|
| "primary failure of eruption" PTH1R | 200 | First 200 results per methodological guidance |
| "impacted canine" "open exposure" "closed exposure" | 200 | First 200 results per methodological guidance |
| "supernumerary tooth" eruption prognosis | 200 | First 200 results per methodological guidance |
| "cleidocranial dysplasia" eruption management | 200 | First 200 results per methodological guidance |

### Citation Tracking of Key Publications (650 records)

| Key Publication | Citation Count | Method |
|---|---|---|
| Suri L, Gagari E, Vastardis H. Delayed tooth eruption: pathogenesis, diagnosis, and treatment. Am J Orthod Dentofacial Orthop. 2004;126(4):432-45. | ~250 | "Cited by" feature; screened for relevance |
| Proffit WR, Vig KW. Primary failure of eruption: a possible cause of posterior open-bite. Am J Orthod. 1981;80(2):173-90. | ~200 | "Cited by" feature; screened for relevance |
| Frazier-Bowers SA, Simmons D, Wright JT, Proffit WR, Ackerman JL. Primary failure of | ~200 | "Cited by" feature; screened for relevance |

| |
|---|
| eruption and PTH1R. Am J Orthod Dentofacial Orthop. 2010;137(2):160.e1-7. |

### Search Strategy Summary

| Database | Records Retrieved | Access Type |
|---|---|---|
| PubMed/MEDLINE | 1,892 | Free access |
| Cochrane Library | 245 | Free access to abstracts; full reviews free after 12 months |
| Google Scholar | 1,450 | Free search access |
| **TOTAL** | **3,587** | |
