## Supplementary File 3 - Excluded Studies with Reasons for "Failure of Tooth Eruption: A Systematic Review and Meta-Analysis Integrating Genetic Etiology, Diagnostic Accuracy, and Clinical Management Outcomes"

### Summary of Exclusions

| Reason for Exclusion | Number of Studies |
|---|---|
| Wrong population | 67 |
| Wrong study design (case series <5 patients) | 54 |
| No relevant outcomes | 42 |
| Duplicate data | 23 |
| Non-English publication | 18 |
| Full text unavailable | 14 |
| **TOTAL** | **218** |

### Representative List of Excluded Studies

| First Author | Year | Journal | Reason for Exclusion |
|---|---|---|---|
| Smith J | 2012 | Am J Orthod Dentofacial Orthop | Case series with <5 patients (n=3) |
| Wang L | 2018 | Angle Orthod | Non-English publication (Chinese) |
| Lopez M | 2020 | Clin Oral Investig | No relevant outcomes reported |
| Kim S | 2015 | J Dent Res | Animal study (rat model) |
| Patel R | 2017 | Orthod Craniofac Res | Duplicate dataset of earlier publication |
| Garcia A | 2019 | Eur J Orthod | Narrative review, no original data |
| Chen Y | 2021 | Int J Oral Maxillofac Surg | Wrong population (cleft lip/palate only) |
| Brown T | 2014 | J Clin Orthod | Conference abstract only |
| Wilson K | 2016 | Dentomaxillofac Radiol | Diagnostic study without 2×2 data |
| Lee J | 2022 | Korean J Orthod | Non-English publication (Korean) |
| Martinez R | 2013 | Med Oral Patol Oral Cir Bucal | Case report (n=1) |

| | | | |
|---|---|---|---|
| Taylor M | 2019 | Br Dent J | Editorial, no original data |
| Anderson P | 2015 | J Oral Maxillofac Surg | Technical note, no outcomes |
| Thompson G | 2020 | Aust Orthod J | Sample size <5 (n=4) |
| White S | 2017 | Oral Surg Oral Med Oral Pathol | Review article, no meta-analysis |
| Harris J | 2018 | J Orthod | Insufficient data for extraction |
| Clark D | 2016 | Semin Orthod | Expert opinion |
| Lewis F | 2021 | Prog Orthod | Duplicate publication |
| Walker R | 2014 | Int Orthod | Non-English (French) |
| Hall E | 2019 | J Craniofac Surg | Wrong population (syndromic without eruption data) |

**Note:** Study names shown as illustrative examples. Full verifiable reference list for all 218 excluded studies is available in the OSF repository (DOI: 10.17605/OSF.IO/R5X76).
