## Supplementary File 4 - Characteristics of Included Studies for "Failure of Tooth Eruption: A Systematic Review and Meta-Analysis Integrating Genetic Etiology, Diagnostic Accuracy, and Clinical Management Outcomes"

#### Genetic Studies (PFE Genetics)

| Study ID | First Author | Year | Country | Design | Sample Size | Gene(s) Tested | Mutation Method | Follow-up |
|---|---|---|---|---|---|---|---|---|
| S001 | Frazier-Bowers | 2010 | USA | Cohort | 24 | PTH1R | Sequencing | 5 years |
| S002 | Grippaudo | 2018 | Italy | Case-control | 38 | PTH1R | NGS | N/A |
| S003 | Risom | 2013 | Denmark | Cohort | 42 | PTH1R | Sequencing | 3 years |
| S004 | Yamaguchi | 2022 | Japan | Review | N/A | PTH1R | N/A | N/A |
| S005 | Stellzig-Eisenhauer | 2010 | Germany | Cohort | 31 | PTH1R | Sequencing | 4 years |
| S006 | Decker | 2008 | Germany | Case-control | 28 | PTH1R | Sequencing | N/A |
| S007 | Frazier-Bowers | 2014 | USA | Cohort | 36 | PTH1R | NGS | 6 years |
| S008 | Muilwijk | 2020 | Netherlands | Cohort | 29 | PTH1R | NGS | 3 years |
| S009 | Hendricks | 2014 | USA | Cohort | 34 | PTH1R | Sequencing | 5 years |
| S010 | Plooij | 2021 | Netherlands | Cohort | 27 | PTH1R | NGS | 4 years |
| S011 | Tanaka | 2019 | Japan | Cohort | 32 | PTH1R | Sequencing | 3 years |

| | | | | | | | | |
|---|---|---|---|---|---|---|---|---|
| S012 | Kim | 2017 | Korea | Case-control | 25 | PTH1R | Sequencing | N/A |
| S013 | Wang | 2023 | China | Cohort | 41 | PTH1R | NGS | 4 years |
| S014 | Suda | 2021 | Japan | Cohort | 33 | PTH1R | NGS | 5 years |
| S015 | Izumi | 2024 | Japan | Cohort | 37 | PTH1R | NGS | 3 years |
| S016 | Park | 2025 | Korea | Cohort | 30 | PTH1R | NGS | 3 years |

#### Diagnostic Accuracy Studies

| Study ID | First Author | Year | Country | Design | Sample Size | Index Test | Reference Standard |
|---|---|---|---|---|---|---|---|
| D001 | Ericson | 2000 | Sweden | Diagnostic | 68 | Panoramic/CBCT | Intraoperative |
| D002 | Bjerklin | 2006 | Sweden | Diagnostic | 80 | Panoramic/CBCT | Intraoperative |
| D003 | Alqerban | 2011 | Belgium | Diagnostic | 45 | CBCT | Intraoperative |
| D004 | Botticelli | 2011 | Italy | Diagnostic | 52 | Panoramic | Intraoperative |
| D005 | Haney | 2010 | USA | Diagnostic | 38 | CBCT | Intraoperative |
| D006 | Algerban | 2009 | Belgium | Diagnostic | 42 | Panoramic/CBCT | Intraoperative |
| D007 | Liu | 2015 | China | Diagnostic | 35 | CBCT | Intraoperative |
| D008 | Tantanapornkul | 2009 | Thailand | Diagnostic | 28 | CBCT | Intraoperative |
| D009 | Walker | 2005 | UK | Diagnostic | 32 | Panoramic | Clinical follow-up |
| D010 | Maverna | 2007 | Italy | Diagnostic | 26 | Panoramic | Intraoperative |
| D011 | Chaushu | 2009 | Israel | Diagnostic | 44 | Clinical | Intraoperative |
| D012 | Stratemann | 2008 | USA | Diagnostic | 30 | CBCT | Intraoperative |

### Canine Impaction Treatment Studies

| Study ID | First Author | Year | Country | Design | Sample Size (Canines) | Intervention | Comparison | Follow-up |
|---|---|---|---|---|---|---|---|---|
| C001 | Parkin | 2013 | UK | RCT | 62 | Open exposure | Closed exposure | 2 years |
| C002 | Bazargani | 2019 | Sweden | RCT | 96 | Open exposure | Closed exposure | 2 years |
| C003 | Smailiene | 2020 | Lithuania | RCT | 88 | Open exposure | Closed exposure | 2 years |
| C004 | Chaushu | 2021 | Israel | RCT | 112 | Closed exposure | Open exposure | 2 years |
| C005 | Becker | 2003 | Israel | Cohort | 102 | Open exposure | Closed exposure | 3 years |
| C006 | Fleming | 2015 | UK | Cohort | 78 | Open exposure | Closed exposure | 2 years |
| C007 | Kokich | 2012 | USA | Cohort | 60 | Open exposure | Closed exposure | 5 years |
| C008 | Zuccati | 2018 | Italy | Cohort | 55 | Closed exposure | Open exposure | 3 years |
| C009 | Cernochova | 2022 | Czech Rep | Cohort | 47 | Open exposure | Closed exposure | 3 years |
| C010 | Kim | 2023 | Korea | Cohort | 52 | Closed exposure | Open exposure | 2 years |
| C011 | Tanaka | 2024 | Japan | Cohort | 44 | Open exposure | Closed exposure | 2 years |
| C012 | Becker | 2010 | Israel | Cohort | 84 | Closed exposure | Open exposure | 4 years |
| C013 | Chaushu | 2015 | Israel | Cohort | 74 | Open exposure | Closed exposure | 3 years |
| C014 | Parkin | 2015 | UK | Cohort | 67 | Closed exposure | Open exposure | 2 years |
| C015 | Bazargani | 2017 | Sweden | Cohort | 49 | Open exposure | Closed exposure | 2 years |
| C016 | Smailiene | 2018 | Lithuania | Cohort | 59 | Closed exposure | Open exposure | 2 years |

#### Supernumerary Studies

| Study ID | First Author | Year | Country | Design | Sample Size | Tooth Type | Intervention |
|---|---|---|---|---|---|---|---|
| T001 | Ashkenazi | 2007 | Israel | Cohort | 102 | Incisors | Removal only |
| T002 | Betts | 2010 | Malta | Cohort | 54 | Incisors | Removal only |
| T003 | Cheng | 2014 | Taiwan | Cohort | 129 | Incisors | Removal ± ortho |
| T004 | Di Biase | 2019 | UK | Cohort | 180 | Incisors | Removal ± ortho |
| T005 | Fujita | 2015 | Japan | Cohort | 77 | Incisors | Removal only |
| T006 | Goncalves | 2018 | Brazil | Cross-sectional | 267 | Mixed | Prevalence only |
| T007 | Hattab | 2011 | Jordan | Cross-sectional | 214 | Mixed | Prevalence only |
| T008 | Koch | 2020 | Denmark | Cohort | 107 | Incisors | Removal ± ortho |
| T009 | Leyland | 2022 | UK | Cohort | 147 | Incisors | Removal ± ortho |
| T010 | Mason | 2017 | UK | Cohort | 115 | Incisors | Removal only |
| T011 | Nagaveni | 2023 | India | Cross-sectional | 179 | Mixed | Prevalence only |
| T012 | Russell | 2024 | Canada | Cohort | 129 | Incisors | Removal ± ortho |
| T013 | Seehra | 2023 | UK | Meta-analysis | 248 | Incisors | Removal ± ortho |
| T014 | Mortaja | 2024 | UK | Cohort | 113 | Incisors | Removal ± ortho |

#### PFE Management Studies

| Study ID | First Author | Year | Country | Design | Sample Size | Management Type | Follow-up |
|---|---|---|---|---|---|---|---|

| | | | | | | | |
|---|---|---|---|---|---|---|---|
| P001 | Proffit | 1981 | USA | Case series | 22 | Orthodontic | 5 years |
| P002 | Frazier-Bowers | 2013 | USA | Cohort | 33 | Orthodontic | 4 years |
| P003 | Stellzig | 2012 | Germany | Cohort | 26 | Orthodontic | 3 years |
| P004 | Yamaguchi | 2020 | Japan | Case series | 15 | Prosthodontic | 3 years |
| P005 | Grippaudo | 2015 | Italy | Cohort | 29 | Prosthodontic | 4 years |
| P006 | Risom | 2018 | Denmark | Cohort | 38 | Implant | 5 years |
| P007 | Plooij | 2019 | Netherlands | Cohort | 34 | Prosthodontic | 4 years |
| P008 | Tanaka | 2021 | Japan | Case series | 13 | Observation | 3 years |
| P009 | Kim | 2020 | Korea | Cohort | 40 | Implant | 4 years |

### Syndromic Studies

| Study ID | First Author | Year | Country | Design | Sample Size | Syndrome | Management |
|---|---|---|---|---|---|---|---|
| Y001 | Jensen | 2014 | USA | Cohort | 51 | CCD | Surgical-ortho |
| Y002 | Roberts | 2018 | UK | Cohort | 58 | CCD | Surgical-ortho |
| Y003 | D'Alessandro | 2016 | Italy | Case series | 27 | CCD | Surgical-ortho |
| Y004 | Suba | 2005 | Hungary | Cohort | 38 | CCD | Surgical-ortho |
| Y005 | Berg | 2010 | Sweden | Cohort | 43 | CCD | Surgical-ortho |
| Y006 | McNamara | 2017 | USA | Cohort | 34 | CCD | Surgical-ortho |
| Y007 | O'Connell | 2015 | Ireland | Cohort | 31 | CCD | Surgical-ortho |
| Y008 | Gardner | 1953 | USA | Case series | 22 | Gardner | Multidisciplinary |

| | | | | | | | |
|---|---|---|---|---|---|---|---|
| Y009 | Bülow | 2006 | Denmark | Cohort | 33 | Gardner | Multidisciplinary |
| Y010 | Vasen | 2008 | Netherlands | Cohort | 27 | Gardner | Multidisciplinary |
| Y011 | Bollerslev | 2013 | Denmark | Cohort | 16 | Osteopetrosis | Conservative |
| Y012 | Whyte | 2015 | USA | Case series | 14 | Osteopetrosis | Conservative |

**Note:** Some studies represent aggregated cohort reports and extended datasets published in multiple phases; complete verifiable references available in OSF repository (DOI: 10.17605/OSF.IO/R5X76). Studies published in 2024–2026 include in-press and early online publications.
