## Supplementary File 5 - Risk of Bias Assessments for "Failure of Tooth Eruption: A Systematic Review and Meta-Analysis Integrating Genetic Etiology, Diagnostic Accuracy, and Clinical Management Outcomes"

### Forest Plots and Funnel Plots

#### List of Figures

| Figure | Title | Description |
|---|---|---|
| Figure S1 | Forest Plot – PTH1R Mutation Prevalence in PFE | 16 studies, 487 patients, $I^2 = 68\%$ |
| Figure S2 | Forest Plot – Diagnostic Sensitivity of Clinical Criteria | Pooled estimates with 95% CI |
| Figure S3 | HSROC Curve – Diagnostic Accuracy of Clinical Criteria | AUC = 0.94 (95% CI: 0.91–0.97) |
| Figure S4 | Forest Plot – Success Rate Difference (Closed vs. Open) | 14 studies, RD 1.6% (-1.8–5.0%) |
| Figure S5 | Forest Plot – Treatment Duration Difference | 7 studies, MD -4.7 months (-7.3 to -2.1) |
| Figure S6 | Forest Plot – Postoperative Pain Difference | 5 studies, MD -1.9 VAS (-2.6 to -1.2) |
| Figure S7 | Forest Plot – Spontaneous Eruption After Supernumerary Removal | 12 studies, range 48–68% |
| Figure S8 | Forest Plot – Prognostic Factors for Supernumerary Success | OR ranges with 95% CI |
| Figure S9 | Funnel Plot – PTH1R Mutation Prevalence | Egger's p = 0.04; trim-and-fill minimal impact |
| Figure S10 | Funnel Plot – Canine Treatment Duration | Egger's p = 0.38; symmetric |
| Figure S11 | Funnel Plot – Canine Postoperative Pain | Egger's p = 0.42; symmetric |
| Figure S12 | Funnel Plot – Supernumerary Spontaneous Eruption | Egger's p = 0.06; some asymmetry |
| Figure S13 | Deeks' Funnel Plot – Diagnostic Accuracy Studies | p = 0.12; no significant asymmetry |
| Figure S14 | Influence Analysis – Leave-One-Out Meta-Analysis | All estimates within CI of pooled |

| | | |
|---|---|---|
| Figure S15 | Subgroup Forest Plot – Familial vs. Sporadic PTH1R | Familial 86% (79–92%), Sporadic 63% (54–71%) |

**Note:** All forest plots and funnel plots are available as separate TIFF files (600 dpi, grayscale) and can be downloaded from the OSF project page: https://osf.io/r5x76/
