## Supplementary File 6 - Forest Plots and Funnel Plots for "Failure of Tooth Eruption: A Systematic Review and Meta-Analysis Integrating Genetic Etiology, Diagnostic Accuracy, and Clinical Management Outcomes"

### GRADE Summary of Findings

#### GRADE Evidence Profile

| Outcome | No. Studies | Study Design | Risk of Bias | Inconsistency | Indirectness | Imprecision | Publication Bias | Certainty |
|---|---|---|---|---|---|---|---|---|
| PTH1R mutation prevalence | 16 | Observational | Serious[1] | Serious[2] | Not serious | Serious[3] | Suspected[4] | ⊕◯◯◯ VERY LOW |
| Diagnostic accuracy (clinical) | 12 | Mixed | Serious[1] | Serious[2] | Not serious | Not serious | Not detected | ⊕⊕◯◯ LOW |
| Diagnostic accuracy (radiographic) | 11 | Mixed | Serious[1] | Moderate[2] | Not serious | Not serious | Not detected | ⊕⊕◯◯ LOW |
| Canine exposure success | 14 | Mixed | Moderate[1] | Moderate[2] | Not serious | Not serious | Not detected | ⊕⊕⊕◯ MODERATE |
| Canine treatment duration | 7 | Mixed | Moderate[1] | Serious[2] | Not serious | Serious[3] | Not detected | ⊕◯◯◯ VERY LOW |
| Canine postoperative pain | 5 | Mixed | Moderate[1] | Moderate[2] | Not serious | Serious[3] | Not detected | ⊕◯◯◯ VERY LOW |

| | | | | | | | | |
|---|---|---|---|---|---|---|---|---|
| Supernumerary spontaneous eruption | 12 | Observational | Serious[1] | Serious[2] | Not serious | Serious[3] | Suspected[4] | ⊕◯◯◯ VERY LOW |
| Supernumerary prognostic factors | 9 | Observational | Serious[1] | Moderate[2] | Not serious | Serious[3] | Not detected | ⊕◯◯◯ VERY LOW |
| PFE orthodontic failure | 7 | Observational | Critical[5] | Serious[2] | Not serious | Serious[3] | Suspected[4] | ⊕◯◯◯ VERY LOW |
| PFE prosthodontic success | 5 | Observational | Critical[5] | Moderate[2] | Not serious | Serious[3] | Suspected[4] | ⊕◯◯◯ VERY LOW |
| Cleidocranial dysplasia | 7 | Observational | Critical[5] | Serious[2] | Not serious | Serious[3] | Not detected | ⊕◯◯◯ VERY LOW |
| Gardner syndrome | 3 | Observational | Critical[5] | Serious[2] | Not serious | Serious[3] | Not detected | ⊕◯◯◯ VERY LOW |
| Osteopetrosis | 2 | Observational | Critical[5] | Serious[2] | Not serious | Very serious[6] | Not detected | ⊕◯◯◯ VERY LOW |

#### GRADE Footnotes

[1] **Downgraded for risk of bias:** Majority of studies had moderate to serious risk of bias due to confounding, selection bias, or lack of blinding.

[2] **Downgraded for inconsistency:** $I^2 > 50\%$ indicates substantial heterogeneity; downgraded one level (serious) or two levels (very serious) based on magnitude.

[3] **Downgraded for imprecision:** Wide confidence intervals crossing clinical decision thresholds or small sample size.

[4] **Downgraded for publication bias:** Funnel plot asymmetry or Egger's test $p < 0.10$.

[5] **Downgraded two levels for risk of bias:** Critical risk of bias due to very small sample sizes, lack of controls, or case series design.

[6] **Downgraded two levels for imprecision:** Very small sample size ($n < 20$) and very wide confidence intervals.

#### GRADE Interpretation

| Symbol | Certainty | Meaning |
|---|---|---|
| ⊕⊕⊕⊕ | HIGH | Further research very unlikely to change confidence |
| ⊕⊕⊕◯ | MODERATE | Further research likely to have important impact |
| ⊕⊕◯◯ | LOW | Further research very likely to have important impact |
| ⊕◯◯◯ | VERY LOW | Any estimate of effect is very uncertain |
