## Supplementary File 7 - GRADE Summary of Findings for "Failure of Tooth Eruption: A Systematic Review and Meta-Analysis Integrating Genetic Etiology, Diagnostic Accuracy, and Clinical Management Outcomes"

### PRISMA 2020 Checklist

| Section and Topic | Item # | Checklist Item | Location (Page) |
|---|---|---|---|
| **TITLE** | | | |
| Title | 1 | Identify the report as a systematic review | 1 |
| **ABSTRACT** | | | |
| Abstract | 2 | See PRISMA 2020 for Abstracts checklist | 2-3 |
| **INTRODUCTION** | | | |
| Rationale | 3 | Describe rationale for review | 4 |
| Objectives | 4 | Provide explicit statement of objectives | 5 |
| **METHODS** | | | |
| Eligibility criteria | 5 | Specify inclusion/exclusion criteria | 6, Table 1 |
| Information sources | 6 | Specify all databases and search dates | 6-7 |
| Search strategy | 7 | Present full search strategy | 7-8, Suppl File 2 |
| Selection process | 8 | Describe selection process | 8 |
| Data collection process | 9 | Describe data extraction methods | 8-9 |
| Data items | 10 | List all outcomes extracted | 9, Suppl File 4 |
| Risk of bias assessment | 11 | Describe risk of bias methods | 9-10 |
| Effect measures | 12 | Specify effect measures | 10 |
| Synthesis methods | 13 | Describe synthesis methods | 10-11 |
| Reporting bias assessment | 14 | Describe publication bias methods | 11 |
| Certainty assessment | 15 | Describe GRADE methods | 11 |
| **RESULTS** | | | |
| Study selection | 16 | Provide study selection results | 12-13, Figure 1 |
| Study characteristics | 17 | Provide study characteristics | 13, Table 2 |
| Risk of bias in studies | 18 | Present risk of bias results | 13-14, Suppl File 5 |

| | | | |
|---|---|---|---|
| Results of individual studies | 19 | Present results for each study | 14-18 |
| Results of syntheses | 20 | Present meta-analysis results | 14-18, Tables 3-8 |
| Reporting biases | 21 | Present publication bias results | Suppl File 6 |
| Certainty of evidence | 22 | Present GRADE results | Suppl File 7 |
| **DISCUSSION** | | | |
| Summary of evidence | 23 | Summarize main findings | 19-20 |
| Limitations | 24 | Discuss limitations | 20 |
| Conclusions | 25 | Provide conclusions | 21-22 |
| **OTHER INFORMATION** | | | |
| Registration | 26 | Provide registration information | 1 |
| Funding | 27 | Describe funding sources | 1 |
| Competing interests | 28 | Declare competing interests | 1 |
| Availability of data | 29 | Provide data availability statement | 1 |

Adapted from: Page MJ, et al. BMJ 2021;372:n71. doi: 10.1136/bmj.n71
