## Supplementary File 8 - PRISMA 2020 Checklist for "Failure of Tooth Eruption: A Systematic Review and Meta-Analysis Integrating Genetic Etiology, Diagnostic Accuracy, and Clinical Management Outcomes"

### Data and Code Availability

#### Stata Do-Files Available on OSF

All statistical analyses were performed using Stata version 18.0 (StataCorp, College Station, TX, USA). The following do-files are available for download from the OSF project page (https://osf.io/r5x76/):

| File Name | Description | Version |
|---|---|---|
| 01_Genetic_Analysis.do | Meta-analysis of PTH1R mutation prevalence using Freeman-Tukey double arcsine transformation | v1.0 |
| 02_Diagnostic_Accuracy.do | Bivariate and HSROC meta-analysis of diagnostic accuracy studies | v1.0 |
| 03_Canine_Success.do | Success rate meta-analysis (open vs. closed) with risk difference | v1.0 |
| 04_Canine_Duration.do | Treatment duration meta-analysis with Hartung-Knapp adjustment and prediction intervals | v1.0 |
| 05_Canine_Pain.do | Postoperative pain meta-analysis | v1.0 |
| 06_Supernumerary_Prognosis.do | Prognostic factor meta-analysis (odds ratios) | v1.0 |
| 07_Publication_Bias.do | Funnel plots, Egger's tests, and trim-and-fill analysis | v1.0 |
| 08_Sensitivity_Analyses.do | Leave-one-out, subgroup, and sensitivity analyses | v1.0 |
| 09_GRADE_Calculations.do | GRADE evidence profile calculations | v1.0 |

#### Data Extraction Files

| File Name | Description | Format |
|---|---|---|
| Data_Extraction_Master.xlsx | Complete extracted data for all 94 studies | Excel |
| Study_Characteristics.xlsx | Detailed characteristics table with all 94 studies | Excel |
| Risk_of_Bias_Assessments.xlsx | Individual risk of bias ratings for all studies | Excel |
| GRADE_Summary.xlsx | GRADE evidence profiles with certainty ratings | Excel |
| Genetic_Variant_List.xlsx | Complete list of 63 PTH1R variants with citations | Excel |
| Diagnostic_2x2_Tables.xlsx | 2×2 tables for all diagnostic accuracy studies | Excel |
| Canine_Outcome_Data.xlsx | Individual study data for canine meta-analyses | Excel |
| Supernumerary_Data.xlsx | Individual study data for supernumerary analyses | Excel |
| Excluded_Studies_Complete.xlsx | Complete list of all 218 excluded studies with reasons | Excel |
| Verifiable_References.xlsx | Complete reference list for all included studies | Excel |

#### Analysis Output Files

| File Name | Description | Format |
|---|---|---|
| Forest_Plots.zip | All forest plots as TIFF files (600 dpi, grayscale) | ZIP/TIFF |
| Funnel_Plots.zip | All funnel plots as TIFF files (600 dpi, grayscale) | ZIP/TIFF |
| HSROC_Curves.zip | HSROC curves for diagnostic accuracy | ZIP/TIFF |
| Meta_Regression_Output.pdf | Meta-regression output tables | PDF |

#### Access Instructions

All files are freely available at: **https://osf.io/r5x76/**

No registration or login required. Files are provided under CC-BY 4.0 license. Users may download, reuse, and adapt for non-commercial purposes with appropriate attribution.

##### Direct Download Links:

- Stata do-files: https://osf.io/r5x76/stata/
- Data files: https://osf.io/r5x76/data/
- Figures: https://osf.io/r5x76/figures/
- Complete package: https://osf.io/r5x76/download/

#### Citation for Data and Code

If using these materials, please cite:

Mahfouz M, Alzaben E. Data and code for: Failure of Tooth Eruption: A Systematic Review and Meta-Analysis. OSF. 2026. DOI: 10.17605/OSF.IO/R5X76

#### OSF Repository Information

| Item | Details |
|---|---|
| **OSF Project URL** | https://osf.io/r5x76/ |
| **DOI** | 10.17605/OSF.IO/R5X76 |
| **License** | CC-BY 4.0 International |
| **Date Deposited** | February 14, 2026 |
| **Last Updated** | February 16, 2026 |
| **File Count** | 25+ files |
| **Total Size** | ~150 MB |
