## Supplementary File 9 - Data and Code Availability for "Failure of Tooth Eruption: A Systematic Review and Meta-Analysis Integrating Genetic Etiology, Diagnostic Accuracy, and Clinical Management Outcomes"

### Risk of Bias Assessments

#### RoB 2.0 (Randomized Controlled Trials)

| Study | Randomization | Deviations | Missing Data | Measurement | Reporting | Overall |
|---|---|---|---|---|---|---|
| Parkin 2013 | Low | Low | Low | Some concerns | Low | Some concerns |
| Bazargani 2019 | Low | Low | Low | Some concerns | Low | Low |
| Smailiene 2020 | Some concerns | Low | Low | Some concerns | Low | Some concerns |
| Chaushu 2021 | Low | Low | Low | Some concerns | Low | Low |
| Becker 2003[1] | High | Some concerns | High | Some concerns | Low | High |
| Fleming 2015 | Low | Low | Some concerns | Some concerns | Low | Some concerns |
| Zuccati 2018 | Low | Low | Low | Low | Low | Low |

[1]Quasi-randomized trial

#### ROBINS-I (Selected Non-Randomized Studies)

| Study | Confounding | Selection | Classification | Deviations | Missing | Measurement | Reporting | Overall |
|---|---|---|---|---|---|---|---|---|

| | | | | | | | | |
|---|---|---|---|---|---|---|---|---|
| Frazier-Bowers 2010 | Moderate | Low | Low | Low | Low | Low | Low | Moderate |
| Grippaudo 2018 | Moderate | Moderate | Low | Low | Low | Low | Low | Moderate |
| Yamaguchi 2022 | Moderate | Low | Low | Low | Low | Low | Moderate | Moderate |
| Renkema 2013 | Moderate | Low | Low | Low | Low | Moderate | Low | Moderate |
| Agudio 2009 | Moderate | Low | Low | Low | Low | Low | Low | Moderate |
| Ashkenazi 2007 | Moderate | Moderate | Low | Low | Moderate | Low | Low | Moderate |
| Ericson 2000 | Moderate | Low | Low | Low | Low | Low | Low | Moderate |
| Becker 2003 | Serious | Moderate | Low | Low | Moderate | Low | Low | Serious |
| Seehra 2023 | Moderate | Low | Low | Low | Low | Low | Low | Moderate |
| Di Biase 2019 | Moderate | Low | Low | Low | Low | Low | Low | Moderate |
| Cheng 2014 | Moderate | Low | Low | Low | Low | Low | Low | Moderate |
| Koch 2020 | Moderate | Low | Low | Low | Low | Low | Low | Moderate |

### QUADAS-2 (Diagnostic Accuracy Studies)

| Study | Patient Selection | Index Test | Reference Standard | Flow/Timing | Overall | Applicability |
|---|---|---|---|---|---|---|
| Ericson 2000 | Low | Low | Unclear | Low | Unclear | Low |
| Bjerklin 2006 | Low | Low | Low | Low | Low | Low |

| | | | | | | |
|---|---|---|---|---|---|---|
| Alqerban 2011 | Low | Low | Low | Low | Low | Low |
| Botticelli 2011 | Low | Unclear | Unclear | Low | Unclear | Low |
| Haney 2010 | Low | Low | Low | Low | Low | Low |
| Algerban 2009 | Low | Low | Low | Low | Low | Low |
| Liu 2015 | Low | Low | Unclear | Low | Unclear | Low |
| Tantanapornkul 2009 | Low | Low | Unclear | Low | Unclear | Low |
| Walker 2005 | Unclear | Unclear | Unclear | Low | Unclear | Unclear |
| Maverna 2007 | Low | Low | Unclear | Low | Unclear | Low |
| Chaushu 2009 | Low | Low | Low | Low | Low | Low |
| Stratemann 2008 | Low | Low | Unclear | Low | Unclear | Low |
